# Genome-wide association study of neuropathic pain phenotypes implicates loci involved in neural cell adhesion, channels, collagen matrix formation and immune regulation

**DOI:** 10.1101/2025.02.04.25320393

**Authors:** Richard Packer, Kayesha Coley, Alexander T Williams, Nick Shrine, Abril G Izquierdo, Jing Chen, Chiara Batini, Mikko Marttila, Balasubramanya S Rao, Raymond Bratty, Frank Dudbridge, William Hennah, Martin D. Tobin

## Abstract

**Background:** Neuropathic pain is a common and debilitating symptom with limited treatment options. Genetic studies, which can provide vital evidence for drug development, have identified only five genome-wide significant signals for neuropathic pain traits. To address this, we performed the largest genome-wide association study (GWAS) to date of all-cause neuropathic pain and neuropathic pain subtypes.

**Methods:** We defined all-cause neuropathic pain and 33 neuropathic pain subtypes using DeepPheWAS software in the UK Biobank, taking advantage of the longitudinal drug prescription data alongside clinical and self-reported records. We performed a GWAS of all-cause neuropathic pain (33,278 cases, 140,134 controls) as our primary analysis and GWASs of neuropathic pain subtypes as secondary analyses. We used eight variant-to-gene criteria to identify putative causal genes.

**Results:** We identified seven independent novel genome-wide associations for neuropathic pain phenotypes which mapped to 22 novel putative causal genes. *NCAM1* was the only gene identified from the primary analysis of all-cause neuropathic pain and met the most variant-to-gene criteria (four) of any identified gene. Of the 21 other genes, *ASCC1, CHST3, C4A/C4B* and *KCNN2* had the most compelling evidence for mechanistic involvement in neuropathic pain.

**Discussion:** We have performed the largest GWAS to date of all-cause neuropathic pain and more than doubled the number of genome-wide significant associations for neuropathic pain traits, identifying putative causal genes. There is strong evidence for the involvement of *NCAM1* in neuropathic pain which merits for further study for drug development.

## Introduction

Neuropathic pain is defined as “pain caused by a lesion or disease of the somatosensory nervous system” (1) and presents a considerable burden on quality of life (2). It is estimated to affect 7-10% of the population (3), with certain at-risk groups, such as people with diabetes, having a much higher prevalence (23-47%). Neuropathic pain syndromes are heterogeneous in their presentation (4) and arise from many primary conditions (5). Drug treatments specific to neuropathic pain have limited effectiveness and significant side effects (6–8). Progress in developing new drugs has been limited (9), and the mechanisms underlying neuropathic pain are less understood than those of nociceptive pain (10).

Genetic studies can improve understanding of disease aetiology and targets for drug development and have potential utility in risk prediction and patient stratification (11,12). Large genetic studies of the broad pain phenotypes of multisite chronic pain (13,14) and pain intensity (15) have generated many genome-wide significant findings (P<5×10). However, the phenotypes studied are not specific to neuropathic pain. Although neuropathic pain is heritable (estimated heritability 37% in twin studies)(16) only a single genome-wide significant association (P<5×10^-8^, mapped to *SLC25A3*) was reported for all-cause neuropathic pain in a study of 4,512 cases (17).

The present study aimed to obtain the largest sample of participants with current or historic all-cause neuropathic pain using novel phenotypes developed in the DeepPheWAS R package (18) in UK Biobank. Unlike previous studies (17), we use primary care data, including prescription records, secondary care data, and study questionnaires, to capture current and historical neuropathic pain.

## Methods

### Phenotype definition

To define our primary phenotype of ‘all-cause neuropathic pain’, we used subject experts to generate a list of neuropathic pain phenotypes; we subdivided this list into those syndromes thought to act centrally (central neuropathic pain) and those acting peripherally (peripheral neuropathic pain). In total, thirty-four neuropathic pain phenotypes were identified: eleven central and 23 peripheral (Supplementary Table 1). Each of the 34 neuropathic pain phenotypes were generated using DeepPheWAS, an open-source platform to facilitate phenome-wide and genome-wide association studies developed in-house (18). DeepPheWAS requires lists of codes and a mapping file describing how to combine those codes to create phenotypes. Briefly, we used lists of codes to define diseases and combined these with a history of neuropathic pain symptoms and/or drug prescriptions for analgesics used explicitly in neuropathic pain.

Symptoms of neuropathic pain were searched for in the Read Code browser (19) and reviewed for inclusion by at least one clinician (Supplementary Tables 2-3). Similarly to previous methods (17), we identified drugs used for neuropathic pain treatment by reviewing well-established guidelines (6,20). We did not exclude participants with depression taking tricyclic medications as depression and chronic pain are commonly co-morbid; we did exclude those with concurrent epilepsy taking anti-epileptics such as pregabalin; we did not use opioids to define cases but did use them to exclude participants as controls. Further details on the method and how each phenotype was defined are in the Supplementary Methods and code lists in Supplementary Tables 2-6.

Finally, we combined ten central neuropathic pain phenotypes and 22 peripheral neuropathic pain phenotypes to form two phenotypes, ‘central neuropathic pain’ and ‘peripheral neuropathic pain’ (Supplementary Table 7). These two phenotypes were then combined to create ‘all-cause neuropathic pain’, giving a total of 37 neuropathic pain phenotypes.

All data was obtained from the UK Biobank study (21) under application 43027. UK Biobank has ethical approval from the UK National Health Service National Research Ethics Service (11/NW/0382), and informed consent was obtained from all participants.

### Genome-wide association

We defined five ancestry groups using k-means clustering and 1000 Genomes Project Phase 3 populations as a reference (22): European (EUR), South Asian (SAS), African (AFR), East Asian (EAS), and American/Hispanic (AMR). For each phenotype-ancestry group combination with at least 200 cases, we performed a genome-wide association study (GWAS) using SAIGEgds (23), using sex, age, array and the first ten principal components as covariates. SNPs with a minor allele count (MAC) below 20 or an INFO score less than or equal to 0.5 were excluded.

We used the LD score intercept (24) to correct for genomic inflation due to possible population stratification. We produced Manhattan and quantile-quantile (QQ) plots for each GWAS. Our primary analysis was the all-cause neuropathic pain phenotype GWAS; secondary analyses were undertaken for all neuropathic pain subtypes.

### Signal selection

In each GWAS we filtered for SNPs that met genome-wide significance (p-value <5×10) and created a 2Mb locus around each SNP. To check for additional independent signals within each locus, we used GCTA 1.94.1 (25,26). We labelled the SNP with the highest posterior probability in each signal as the sentinel SNP, and we annotated the nearest gene using ANNOVAR (27). We calculated a 99% credible set for each signal using the Wakefield method (28), with a prior *W* of 0.04. For each signal, we produced a region plot annotated with the credible sets using LocusZoom (29).

To fine-map signals within the HLA region (chr6:29,607,078-33,267,103 (b37)) to a specific HLA allele or amino acid change, we used imputed HLA amino acid changes and classical alleles. Then, we repeated the association testing as described above. To test whether an imputed HLA allele was driving the association, we used GCTA cojo-cond command to condition the associations on all the available HLA variants.

### Assessing the novelty of sentinel variants

We assessed our sentinel variants as novel if LD was r^2^ <0.2 with any of the five previously identified genome-wide significant sentinels from GWAS of neuropathic pain traits identified from Reyes-Gibby CC et al.(30) and Veluchamy et al. (17).

### Identifying putative causal genes

We combined evidence from eight sources: (i) the nearest gene to each sentinel variant; (ii) the gene with the highest Polygenic Priority Score (PoPS) (31), an approach that assumes genes that are close to sentinel variants and that share functional characteristics are most likely to be causal; colocalisation of (iii) expression quantitative trait loci (eQTLs) and (iv) protein quantitative trait loci (pQTLs) with pain GWAS signals; (v) proximity to a gene for pain-related Mendelian disease (±500kb); (vi) identification of a rare variant associated with pain phenotypes using results from exome-wide association studies and (vii) gene-based testing in UK Biobank; and (viii) proximity to a human ortholog of a mouse knockout gene with a pain-related phenotype (±500kb), (see Supplementary Methods). Where a credible set was defined in non-EUR populations, we did not perform colocalization analysis but simply queried eQTL and pQTL resources instead.

For any gene identified through these methods we undertook a literature search using OMIM (32), National Centre for BioTechnology information Gene (33),GWAS Catalog (34), the Protein Atlas (35) and Google Scholar to identify gene function and search for existing associations with pain phenotypes. References from initial findings were used to find any additional relevant studies.

We characterised genes as novel for neuropathic pain if they had not been previously reported in Reyes-Gibby CC et al.(30) and Veluchamy et al. (17).

### Comparison with reported genetic signals for pain intensity

We identified independent genome-wide significant variants reported in a GWAS of pain intensity (15) and in two GWAS of multisite chronic pain (13,14), and performed a Binomial test to check for enrichment in our GWAS results.

### Association with other phenotypes

We undertook a PheWAS for each non-HLA sentinel variant in 2303 phenotypes using DeepPheWAS (18) and reported any association with a false discovery rate (FDR) <1%. Results were not reported when the only association was with the GWAS trait. Results were available for EUR, AFR, SAS and AMR ancestries. To supplement DeepPheWAS we also searched Open Targets Genetics Portal (version 8; accessed 27^th^ September 2024 (36)) and the FinnGen browser (DF11 – accessed 27^th^ September 2024) (37) for each of the sentinel variants.

## Results

### Neuropathic pain phenotypes

For the GWAS of the primary phenotype, all-cause neuropathic pain, 34,278 cases and 140,134 controls were available in Europeans; African, South Asian and American/Hispanic ancestry subgroups had 359, 550, and 342 all-cause neuropathic pain cases, respectively. Secondary GWAS for neuropathic pain subtypes were feasible (>200 cases) in 28 of 180 phenotype-ancestry group combinations (Supplementary Table 8).

### Associations with neuropathic pain traits

The primary analysis of all-cause neuropathic pain showed a single independent genome-wide significant locus (*NCAM1*) in Europeans and a single locus (*MIR548XHG*) in Africans (Table 1); neither association has been previously reported for neuropathic pain, although a variant, rs1940720, in weak LD (r^2^ = 0.38) with our *NCAM1* sentinel, rs12277922, previously showed association with back pain (Supplementary Table 9)(38). rs12277922 associations with all tested neuropathic pain traits in Europeans (Supplementary Tables 7) are shown in Figure 1.

**Figure 1:**
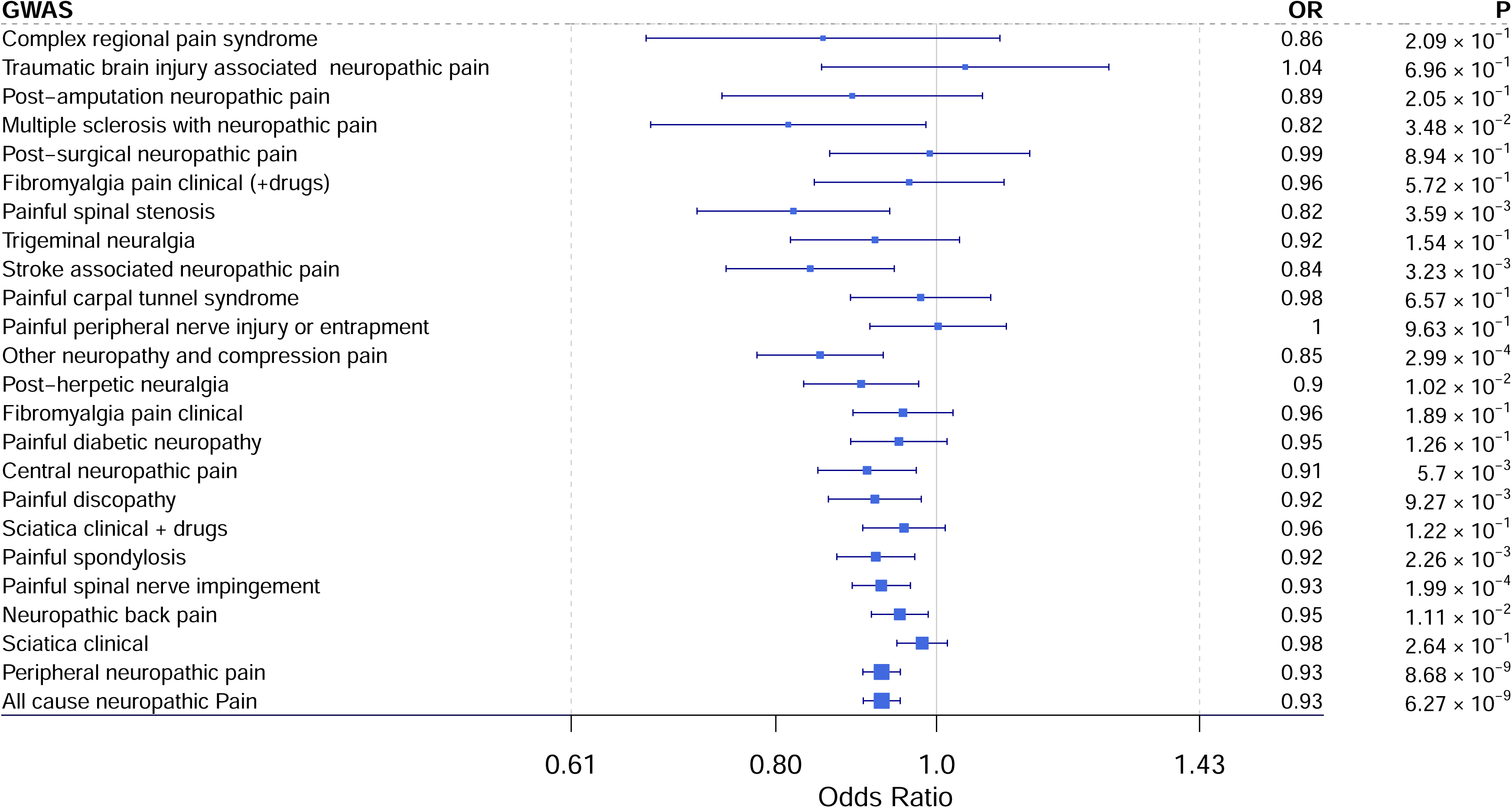
Forest plot of NCAM1 sentinel variant rs12277922 associations with neuropathic pain phenotypes in EUR sample (OR = Odds ratio)

**Table 1:** Genome-wide significant loci with sentinel variants from GWAS of neuropathic pain phenotypes. Chr = chromosome, BP = base pair, MAF = Minor allele frequency, CI = confidence interval, ª=Loci was also found in peripheral neuropathic pain GWAS in the same ancestry, EUR = European ancestry, AFR = African ancestry

| GWAS (Ancestry) | N-cases /<br>N-controls | Chr:Position<br>(GRCh37) | RSID | Locus (BP distance<br>from gene) | Coded/non-<br>coded allele | MAF (minor<br>allele) | OR (95 CI) | P | Genes mapped using<br>variant-to-gene criteria |
| --- | --- | --- | --- | --- | --- | --- | --- | --- | --- |
| All-cause neuropathic<br>Pain<sup>a</sup> (EUR) | 34278 /<br>140134 | 11:112917378 | rs12277922 | *NCAM1* (intronic) | C/T | 20% (T) | 0.93 (0.9-0.95) | 6.27x10<sup>-9</sup> | *NCAM1* |
| Sciatica clinical (EUR) | 10600 /<br>181898 | 10:73933699 | rs11000202 | *ASCC1* (intronic) | A/G | 48% (A) | 0.93 (0.9-0.95) | 4.65x10<sup>-8</sup> | *ASCC1, CDH23, DDIT4,*<br>*MICU1, PSAP, ANAPC16,*<br>*CHST3* |
| Sciatica clinical + drugs<br>(EUR) | 5089 /<br>17185 | 1:64613556 | rs19269295<br>9 | *ROR1* (intronic) | A/G | 0.15% (G) | 0.17 (0.09-0.31) | 1.71x10<sup>-8</sup> | *ROR1* |
| Fibromyalgia clinical<br>(EUR) | 4773 /<br>211196 | 1:192923542 | rs946556 | *RGS2-AS1* (6155) | G/A | 0.1% (A) | 0.04 (0.01-0.12) | 2.03x10<sup>-9</sup> | *RGS2, CDC73, CFHR1* |
| Central neuropathic<br>pain (EUR) | 2801 /<br>148886 | 6:32460831 | rs77078150 | *HLA-DRB5* (24323) | G/T | 17% (G) | 1.24 (1.16-1.34) | 5.84x10<sup>-9</sup> | *C4A/C4B, HLA-DQA2,*<br>*HLA-DRB5, TNXB, AGER,*<br>*BTNL2, BRD2* |
| Post-surgical<br>neuropathic pain EUR) | 695 /<br>12905 | 5:113421943 | rs76420936 | *KCNN2* (276073) | G/A | 1% (A) | 0.19 (0.11-0.34) | 3.19x10<sup>-8</sup> | *KCNN2* |
| All-cause neuropathic<br>Pain<sup>a</sup> (AFR) | 550 / 1467 | 21:20113911 | rs2825061 | *MIR548XHG*<br>(intronic) | C/T | 1% (T) | 0.38 (0.27-0.54) | 3.29x10<sup>-8</sup> | *MIR54XHG* |

Secondary GWAS for pain subtypes in Europeans identified five further independent loci, none of which were previously reported. These were for Sciatica clinical (*ASCC1*), Sciatica clinical with drugs (*ROR1*), Fibromyalgia clinical (*RGS2-AS1*), Central neuropathic pain (*HLA-DRB5*) and Post-surgical neuropathic pain (*KCNN2*) (Table 1). Given one locus was in the HLA region, we also imputed HLA alleles, although as none were associated with Central neuropathic pain at genome-wide significance, we retained the sentinel SNP association and associated credible set for subsequent analyses. See Supplementary Figures 1-26 for QQ, Manhattan, and region plots for all genome-wide significant signals.

Neither the single genome-wide association (P1.3×10^-8^, *SLC25A3*) previously reported for all-cause neuropathic pain in a study of 4,512 cases (17), nor four previously reported associations (*SNX8*, *PCP2*, *KNG1*, *RORA*) in only 130 head and neck cancer pain cases (30) replicated in our study (P<0.01).

### Variant to gene mapping and PheWAS

We mapped 22 genes across the seven independent signals; five genes (*C4A/C4B, ROR1, KCNN2, RGS2*) fulfilled two variant-to-gene criteria, one gene (*ASCC1*) three criteria and a single gene, *NCAM1*, identified in our primary analysis of all-cause neuropathic pain in Europeans fulfilled four criteria (Table 2, Supplementary Tables 10-15). None of these genes have been previously implicated in genetic studies of neuropathic pain. We mapped no genes from the functional annotation of credible sets.

**Table 2:** Variant to gene (V2G) mapping results for all independent signals.

| gene | Number of V2G criteria | Variant-to-gene (V2G) criteria met | GWAS trait | Ancestry |
| --- | --- | --- | --- | --- |
| *NCAM1* | 4 | eQTL, pQTL, PoPs, nearest gene | all-cause neuropathic pain | EUR |
| *ASCC1* | 3 | eQTL, nearest gene, rare variant | sciatica clinical | EUR |
| *C4A/C4B* | 2 | rare variant, gene-based test | central neuropathic pain | EUR |
| *RGS2* | 2 | PoPs, nearest gene | fibromyalgia clinical | EUR |
| *KCNN2* | 2 | PoPs, nearest gene | post-surgical neuropathic pain | EUR |
| *CDH23* | 2 | eQTL, rare variant | sciatica clinical | EUR |
| *ROR1* | 2 | PoPs, nearest gene | sciatica drugs | EUR |
| *MIR54XHG* | 1 | nearest gene | all-cause neuropathic pain | AFR |
| *HLA-DQA2* | 1 | PoPs | central neuropathic pain | EUR |
| *HLA-DRB5* | 1 | nearest gene | central neuropathic pain | EUR |
| *TNXB* | 1 | rare disease | central neuropathic pain | EUR |
| *AGER* | 1 | mouse KO | central neuropathic pain | EUR |
| *BTNL2* | 1 | rare variant | central neuropathic pain | EUR |
| *BRD2* | 1 | rare variant | central neuropathic pain | EUR |
| *CDC73* | 1 | rare disease | fibromyalgia clinical | EUR |
| *CFHR1* | 1 | pQTL | fibromyalgia clinical | EUR |
| *CHODL* | 1 | PoPs | peripheral neuropathic pain | AFR |
| *DDIT4* | 1 | PoPs | sciatica clinical | EUR |
| *MICU1* | 1 | rare disease | sciatica clinical | EUR |
| *PSAP* | 1 | rare disease | sciatica clinical | EUR |
| *ANAPC16* | 1 | eQTL | sciatica clinical | EUR |
| *CHST3* | 1 | Gene-based test | sciatica clinical | EUR |

### NCAM1 Locus

Results from colocalised eQTL and pQTL showed that lower risk of all-cause neuropathic pain was aligned with higher expression of *NCAM1* in blood and higher circulating NCAM1 protein levels (Supplementary Tables 10 and 11). The nearest gene, *NCAM1*, encodes the protein Neural cell adhesion molecule 1, involved in neural regeneration, cell-to-cell adhesion and signal transduction (39,40). *NCAM1* is expressed within the central nervous system, peripheral nerves, heart and muscles (35). In humans, NCAM1 has been identified by ELISA in serum samples taken from patients with inflammatory neuropathies (41), and patients with Charcot Marie Tooth syndrome (42). In rat models of neuropathic pain, the analgesic effect of glial cell line-derived neurotrophic factor was mediated through NCAM1 signalling, and *NCAM1* was expressed in intrinsic neurons in the spinal dorsal horn and dorsal root ganglion (43). In mouse models of peripheral nerve injury, *NCAM1* was implicated in synaptic plasticity, structural and behavioural changes after injury and pain sensitisation (44).

In DeepPheWAS, the NCAM1 sentinel, rs12277922 was associated (FDR<1%) with 12 traits, all in Europeans. The allele (T) associated with an increased risk of all-cause neuropathic pain was associated with an increased risk of ever smoking, comparatively smaller body size, higher risk of back pain, ‘other mental disorders’, and extrapyramidal disease. The risk allele (T) was also associated with lower levels of IGF-1, higher levels of gamma glutamyl transferase and apolipoprotein and lower forced expiratory volume in 1 second over forced vital capacity ratio (Supplementary Table 16). Results for the same variant (rs12277922) in FinnGen showed that the risk allele (T) was associated with an increased risk of dorsalgia, joint pain, depression and use of antidepressant medications (Supplementary Table 17). A search of OpenTargets GWAS lead variants in LD with our lead variant (rs12277922) showed an association between higher risk of all-cause neuropathic pain and higher risk of neurological diseases, lower age at first sexual intercourse, higher levels of CD16-CD56 natural killer cells, lower intelligence and lower serum levels of NCAM1 (Supplementary Table 19).

### ASCC1 locus

The signal for sciatica phenotype mapped to seven genes: *ASCC1, CHST3, CDH23, PSAP*, *ANAPC16*, *MICU1* and *DDIT4. ASCC1* fulfilled three variant-to-gene criteria. It was the nearest gene in the *ASCC1* locus for the sciatica clinical GWAS, colocalised with an eQTL signal and had a rare variant association with a pain phenotype (Table 2). The sentinel variant, rs11000202, the risk allele for sciatica (G), was associated with increased expression of *ASCC1* in blood (Supplementary Table 10). The rare variant association was found for multisite chronic pain and was intronic (Supplementary Table 14). ASCC1 is a tetrametric nuclear-activating signal integrator 1 (ASC-1) complex subunit. Dysfunction in this receptor complex has been shown in mouse models, zebra fish embryos and cell lines to be critical for a functional neuromuscular junction (45). In humans in vivo, loss of function mutations in *ASCC1* cause manifestations of spinal muscular atrophy with congenital bone fractures (46), a further in vivo mutation overexpressed truncated ASCC1, presumed causal for congenital myopathy (47).

*CHST3* was identified in a gene-based association test for back pain (P=0.00019, Supplementary Table 15). *CHST3* has not previously been associated with neuropathic pain but has shown associations in GWAS of other non-neuropathic pain susceptibility traits (48); lumbar disc degeneration (49,50), osteoarthritis (51), intervertebral disc disorder (52) and back pain (38). *CHST3* is expressed widely (35) and encodes sulfation of chondroitin into chondroitin sulfate proteoglycans, which have been investigated as a potential therapeutic compound for osteoarthritis (53) as well as being implicated as important regulators of nerve regeneration (54). Mendelian disorders of *CHST3* are characterised by spondyloepiphyseal dysplasia and congenital joint dislocations (55–59).

*CDH23* was prioritised by a colocalised eQTL (Supplementary Table 10) and by a single rare variant association (Supplementary Table 15). The sentinel variant, rs11000202, the protective allele (A) for sciatica, was associated with reduced expression of *CDH23* in blood. The rare variant was associated with fibromyalgia and was intronic. In humans, *CDH23* has a well-characterised mendelian disorder, Usher Syndrome 1D, which presents with progressive hearing and vision loss (60). *CDH23* has not previously been associated with neuropathic pain, but was identified at nominal significance (P=6.25×10^-5^) in a gene-based analysis of an at-home cold suppressor test (61).

*PSAP* was prioritised given its association with the rare lysosomal disorder Metachromatic leukodystrophy (Supplementary Table 13), characterised by myelin sheath breakdown and often neuropathy (62). *ANAPC16*, *MICU1*, *DDIT4*, and *ROR1* identified for the related sciatica drugs phenotype have no previous evidence of mechanistic association with pain.

For the ASCC1 locus sentinel variant, rs11000202, results from DeepPheWAS showed two associations (excluding discovery phenotype) at FDR significance (FDR <0.01); the risk allele for higher risk of sciatica (G) was associated with being smaller at age 10 and higher risk of back pain (Supplementary Table 16). Within FinnGen, the same variant was associated with a lower risk of arthropathies, including gonarthrosis (P=7.3×10), but higher risk of prolapsed discs (P=7.3×10) and dorsalgia (P=1.3×10) (Supplementary Table 17).

### KCNN2 locus

*KCNN2,* the only gene implicated in the post-surgical neuropathic pain GWAS, by PoPs and nearest gene criteria (Table 1, Table 2), is a calcium-activated potassium channel. In humans, it is implicated in myoclonic-dystonia (63). In mouse models, deletion/blocking of *KCNN2* consistently reports a tremor phenotype (64–66), which has also been reported with novel mutations in humans (67). Experiments in mouse models showed a plausible link between *KCNN2* and neuropathic pain (68). Here, the sciatic nerve damage was damaged, which reduced parvalbumin expression in parvalbumin-expressing cells (PV-cells) located on the dorsal horn of the spinal cord. PV-cells act as inhibitors to onward nerve transmission through continuous nerve firing. Parvalbumin within these cells acts as a buffer for intra-cellular calcium, and reduced expression leads to raised intracellular calcium and activation of KCNN2. This leads to adaptive firing, which reduces the inhibitory function of PV cells and mechanical allodynia in mouse models. Selective blocking of KCNN2 led to resolving mechanical allodynia. Within FinnGen, the sentinel variant for the *KCNN2* locus, rs76420936 risk allele A, showed association with increased risk of dorsalgia (P=9×10^-5^), lower back pain (P=6×10^-5^), sciatica (P=1×10^-4^) and general pain (P=3×10^-4^) (Supplementary Table 17).

### RGS2 locus

The signal for fibromyalgia phenotype mapped to three genes. *RGS2* was prioritised with two variant-to-gene criteria (PoPs, nearest gene, Table 2) and is a regulator of G-protein activity. In mouse models, deletion of *RGS2* induces hypertension and renovascular abnormalities (69). G-protein signalling has been implicated in nociceptive and anti-nociceptive neuronal signalling; G-protein activity may be analgesic or pain-promoting depending on the cell and location (70). *CFHR1* and *CDC73* had no previous evidence of mechanistic association with pain.

### HLA-DRB5 locus

The signal for central neuropathic pain phenotype located in the HLA region mapped to seven genes, *C4A/C4B, TNXB, BRD2, BTNL2, HLA-DQA2*, *HLA-DRB5* and *AGER,* none of which have been associated with neuropathic pain via GWAS studies previously. *C4A/C4B* were identified at nominal significance in rare variant and gene-based tests. They are part of the complement system of innate immunity, with antimicrobial action and a role in inflammatory regulation (71). The complement pathway has been implicated in mouse models of neuropathic pain (72–74) studying pro-inflammatory C3 and C5 subunits (73,74), one caveat being that C4 has lower catalytic activity in mouse than in human (75). *TNXB* belongs to the tenascin family of extracellular matrix glycoproteins relevant to wound healing; a variant of *TNXB* causes Ehlers-Danlos syndrome (76,77) which is associated with chronic neuropathic pain (78). In mouse models *TNXB* has been reported to function through dermal fibroblast collagen deposition (79) and isolated hypersensitivity of Aδ- and Aβ-fibers upregulated pain signals (80). *TNXB* variant associations were reported in a GWAS of back pain (38). *BRD2* is a transcription regulator and part of the BET family of proteins; BRD4 protein blockers have a therapeutic effect in mouse models of osteoarthritis (81) and neuropathic pain (82–84) through inhibition of the inflammatory response. *BTNL2*, is an immunoregulator (85) and has been implicated in a single GWAS of osteoarthritis in a Japanese population (86). *HLA-DQA2*, *HLA-DRB5* or *AGER* have no previous evidence of mechanistic association with pain.

### MIR548XHG locus

The signal from all-cause neuropathic pain in African ancestry was mapped only by the nearest gene criterion to *MIR548XHG*. This gene is affiliated with long noncoding RNAs and investigated as a prognostic marker for lung cancer and hepatic cancer (87,88). Within the same cytogenic band (21q21.1), approximately 2Mb upstream of *MIR548XHG* is *NCAM2,* a paralog of *NCAM1*. Using Olink proteomic data in the UK Biobank, we found no association of our sentinel variant, rs2825061, with NCAM2 levels in African (P=0.69) or European (P=0.28) samples.

### Comparison with reported genetic signals for pain intensity

Of 259 sentinel variants previously reported as reaching genome-wide significance in a GWAS of pain intensity (15), 93 showed an association with neuropathic pain in our GWAS, more than expected by chance (Binomial P=2.2×10^-16^). Of 41 sentinels previously reported as reaching genome-wide significance in a GWAS of multisite chronic pain (13,14), 11 showed association with neuropathic pain in our GWAS, more than expected by chance (Binomial P=3.8×10^-6^). 41 of the 93 sentinels for pain intensity and all 11 of the multisite chronic pain sentinels also showed consistent direction of effect to our study.

## Discussion

We performed the largest GWAS to date of all-cause neuropathic pain, identifying a single genome-wide significant signal mapped with four variant-to-gene criteria (nearest gene, PoPs, eQTL, pQTL) to a single putative causal gene, *NCAM1*. The sentinel variant and gene have not previously been associated with neuropathic pain in GWAS. In an independent population, the same *NCAM1* variant was associated with non-neuropathic pain phenotypes, corroborating a role of *NCAM1* in pain. *NCAM1* has plausible mechanisms for pain regulation in mouse models (43,44) and elevated levels of NCAM1 have been found in serum samples from those with chronic inflammatory neuropathies (41,42). Although *NCAM1* is expressed widely in central and peripheral nervous system (35) statistical colocalisation of *NCAM1* eQTL and GWAS signals was only shown in blood, possibly explained by the smaller sample size and power for eQTL studies in non-blood tissues (eQTL gen N=31,684, neural tissues in GTEX N ranged from 181-300). In our PheWAS, the risk allele for neuropathic pain of the *NCAM1* sentinel variant was additionally associated with increased risk of back pain, mental health disorders and decreased lung function.

In our GWASs of neuropathic pain subtypes, we found six additional signals at genome-wide significance and mapped these to 21 putative causal genes. All six sentinel variants and 21 genes have not previously been implicated in any neuropathic pain trait. Together with our primary analysis of all-cause neuropathic pain, this represents a more than doubling of the five existing signals for neuropathic pain traits.

The *ASCC1* locus we identified for sciatica mapped to seven genes. *CHST3* identified via association in in collapsing gene-based testing of the related phenotype fibromyalgia, had the most substantial previous evidence of implication in non-neuropathic pain of the seven prioritised genes and has been implicated in previous GWAS of pain susceptibility traits (48), lumbar disc degeneration (49,50), osteoarthritis (51), intervertebral disc disorder (52) and back pain (38). The PheWAS of the sentinel variant showed minimal pleiotropy, showing the risk of sciatica was associated with being comparatively smaller as a child and increased risk of back pain. Functionally, *CHST3* produces chondroitin sulphate, an important chemical in cartilage composition. Mendelian disorders of *CHST3* result in severe joint dysfunction.

The single genome-wide significant finding in our GWAS of central neuropathic pain was within the HLA region. We were unable to map to a HLA allele, and our variant-to-gene mapping identified evidence for seven genes. *C4a/C4b* was identified through rare-variant and gene-based testing, is part of the complement system and has shown involvement with neuropathic pain models in mice (72–74), but not previously in humans. More research with human models of the complement system could better assess C4 as a drug target for pain.

We identified a single gene in our GWAS of post-surgical neuropathic pain, *KCNN2*, which has a mechanistically plausible function in de-regulating inhibitory neurons (68) in mice. The sentinel variant is rare (MAF 1%), and the GWAS had a small sample size (695 cases), leading to a greater potential for a false positive. Further, pharmacological blockage of this gene is challenging; loss of function mutations in *KCNN2* produce a consistent tremor phenotype in mice (64–66) and have shown to myoclonic-dystonia and tremor phenotypes in humans (67).

Finally, our GWAS of fibromyalgia identified three genes *RGS2, CFHR1* and *CDC73*; our literature review did not identify evidence implicating these genes in pain.

Our approach of exploiting the extensive linked electronic health care records in UK Biobank using DeepPheWAS led to us gathering the largest sample of all-cause neuropathic pain, and by analysing the subtypes of pain that make up our all-cause phenotype we have been able to generate additional insights. Further, we provide a comprehensive account of code lists used and how we combined these lists to generate phenotypes. We used publicly available software to do so, allowing replication and use of our phenotype definition to be used by others. Challenges in phenotyping neuropathic pain have been well documented and questionnaires designed to distinguish somatic from neuropathic pain has been recommended for genetic studies (3). When defining our phenotypes, we had to make many decisions about including or excluding codes and how to combine them. For example, unlike the only previous published study of all-cause neuropathic pain (17) we chose to include tricyclic antidepressants in the definition of neuropathic pain drugs and did not exclude those with a record of depression. We did this as pain and depression commonly occur together and may share common mechanisms (89). However, this may have led to misclassification of some individuals as having neuropathic pain if they solely had depression. Like many cohort-based studies, we lacked power in non-European ancestries.

## Conclusion

Overall, these results demonstrate the viability of using electronic health care records to define neuropathic pain phenotypes, which enables the study of their genetic determinants and shows strong evidence for the involvement of *NCAM1* in neuropathic pain risk. Analysis of subtypes of neuropathic pain showed good evidence for the *ASCC1* locus, adding to the historic GWAS association of back, lumbar disc and osteoarthritic pain, although mechanistically, this may be driven by joint dysfunction. These loci warrant further investigation for potential drug development.

## Data Availability

All data was obtained from the UK Biobank study. Code to reproduce the pain phenotypes can be obtained using the DeepPheWAS R package (https://github.com/Richard-Packer/DeepPheWAS)

## Acknowledgements

The research was partially supported by the NIHR Leicester Biomedical Research Centre and through an NIHR Senior Investigator Award to M.D.T.; views expressed are those of the author(s) and not necessarily those of the NHS, the NIHR or the Department of Health. The funders had no role in the design of the study. For the purpose of open access, the author has applied a CC BY public copyright licence to any Author Accepted Manuscript version arising from this submission. C.B. was supported by a UKRI Innovation Fellowship at Health Data Research UK (MR/S003762/1).

MDT, RP, ATW, FD and NS received funding from Orion Pharma within the scope of the submitted work. WH, and MM are salaried employees of Orion Pharma within the scope of the submitted work. RB and BR were previously salaried employees of Orion within the scope of this work; BR is now a salaried employee of AstraZeneca. We thank all participants and staff who have contributed their time to this study. Our analyses used the ALICE and SPECTRE High Performance Computing Facilities at the University of Leicester. We are grateful to the staff who develop and maintain the GWAS Catalog, and also acknowledge the participants and investigators of the FinnGen study.

## Supplementary table legend

- **Supplementary Table 1**: Central and peripheral neuropathic pain phenotypes.
- **Supplementary Table 2:** Concepts listed with Concept name matching phenotype formula, broad type of concept and written description. Code lists for Clinical concepts in table S3, medication concepts S4 and S5. Field ID concept field ID and value extracted in table S6. Composite concept formula is shown in supplementary materials.
- **Supplementary Table 3:** Code list for all clinical concepts to search filter by ‘Concept name’, V3 = Read Version 3 codes, MD = Mortality data, cancer = cancer registry, ICD9 = Internation disease classification version 9, ICD10 = International disease classification version 10, SR = self-report non-cancer diagnosis, SROP = Self-reported operations, OPCS = OPCS Classification of Interventions and Procedures version 4, 1 = included, 0 = excluded.
- **Supplementary Table 4:** Read V2 code list for all medication concepts, to search filter by ‘Concept name’, 1 = included, 0 = excluded.
- **Supplementary Table 5:** Unique search term used for all medication concepts to search filter by ‘Concept name’.
- **Supplementary Table 6:** Field ID and values extracted for field_ID concepts to search filter by ‘phenotype name’.
- **Supplementary Table 7**: Phenotypes combined to form central neuropathic pain phenotype and peripheral neuropathic pain phenotype.
- **Supplementary Table 8:** Summary of case and control numbers for Neuropathic pain phenotypes created by DeepPheWAS using UK Biobank data across five ancestry groups. Those highlighted in orange met the required 200 cases for progression to genome-wide association analysis, those in grey did not. Phenotypes in bold are formed by combining the cases of other phenotypes. **\* = Cases for phenotype included in P2000.1 Central neuropathic pain. ** = Cases for phenotype included in P2000.3 Peripheral neuropathic pain. ^ = Cases for phenotype included in P2000 All-cause neuropathic pain.**
- **Supplementary Table 9:** Result of queries for GWAS Catalog findings in neuropathic pain GWAS discovery loci. OR=odds ratio, CI=confidence interval, R2=squared correlation (linkage disequilibrium) with sentinel variant for each GWAS, *result altered from GWAS Catalog based upon search of summary statistics. Note original results for rs1940720 and rs28732174 from (213) were removed as a search of the available summary statistics showed the lowest P values (identified in the meta-analysis) were 8.85×10-7 and 7.95×10-5, respectively. rs2219837 and rs3180 from (212) were removed as a search of summary statistics showed P values of 9.8×10-1 and 9.0×10-2, respectively.
- **Supplementary Table 10**: eQTL co-localising SNP-gene expressions, all associations are in eQTL gen blood. Pval.eqtl - p-value for eQTL and SNP, se = standard error.
- **Supplementary Table 11**: pQTL colocalisation results, direction of effect is aligned with the sentinel variant for each GWAS as reported in Table 1, H4 = posterior probability, DOE = direction of effect, UKB = UK Biobank, * = colocalised with coloc.abf and no SuSiE.
- **Supplementary Table 12:** Genes prioritised by PoPs method for variant to gene mapping.
- **Supplementary Table 13:** Results for rare disease search within 500Kb of sentinal variants for variant to gene mapping.
- **Supplementary Table 14:** Rare variant exome wide association results for variant to gene mapping, genomic coordinates using hg38 build. Chr=chromosome.
- **Supplementary Table 15:** Collapsed gene testing results for variant to gene mapping.
- **Supplementary Table 16**: DeepPheWAS result for sentinel variants. FDR=false discovery rate, OR=odds ratio, U95=upper 95% confidence interval, L95=lower 95% confidence interval, MAC=minor allele count, N_ID,=number of unique IDs in analysis, use variant or Locus to filter by locus.
- **Supplementary Table 17:** Query results of sentinel variants in FinnGen. Only results with p-value <0.05 are included. To search per signal filter locus. Pval = p-value, maf = minor allele frequency, maf_cases = minor allele frequency in cases, n_cases = number of cases, n_controls = number of controls, pip = posterior inclusion probability.
- **Supplementary Table 18:** Query results of sentinel variants in OpenTargets genetics GWAS lead variants. Only results with p-value <5×10-8 are included. Beta reported is aligned to the protective allele from our GWAS. To search per signal filter locus. PMID = pubmed ID, Study N = Number in the study, LD = linkage disequilibrium between lead variant and pain GWAS variant.

## References

1. Terminology | International Association for the Study of Pain [Internet]. International Association for the Study of Pain (IASP). [cited 2023 Mar 13]. Available from: https://www.iasp-pain.org/resources/terminology/

2. Jensen MP, Chodroff MJ, Dworkin RH. The impact of neuropathic pain on health-related quality of life: Review and implications. Neurology. 2007 Apr 10;68(15):1178–82.

3. van Hecke O, Kamerman PR, Attal N, Baron R, Bjornsdottir G, Bennett DLH, et al. Neuropathic pain phenotyping by international consensus (NeuroPPIC) for genetic studies: a NeuPSIG systematic review, Delphi survey, and expert panel recommendations. Pain. 2015 Nov;156(11):2337–53.

4. Bouhassira D. Neuropathic pain: Definition, assessment and epidemiology. Revue Neurologique. 2019 Jan;175(1–2):16–25.

5. Smith PA. Neuropathic pain; what we know and what we should do about it. Front Pain Res. 2023 Sep 22;4:1220034.

6. Finnerup NB, Attal N, Haroutounian S, McNicol E, Baron R, Dworkin RH, et al. Pharmacotherapy for neuropathic pain in adults: a systematic review and meta-analysis. The Lancet Neurology. 2015 Feb;14(2):162–73.

7. Additional information | Neuropathic pain in adults: pharmacological management in non-specialist settings | Guidance | NICE [Internet]. NICE; 2013 [cited 2023 Mar 13]. Available from: https://www.nice.org.uk/guidance/cg173/chapter/Additional-information

8. Amitriptyline hydrochloride | Drugs | BNF content published by NICE [Internet]. [cited 2023 Mar 27]. Available from: https://bnf.nice.org.uk/drugs/amitriptyline-hydrochloride/

9. Price TJ, Basbaum AI, Bresnahan J, Chambers JF, De Koninck Y, Edwards RR, et al. Transition to chronic pain: opportunities for novel therapeutics. Nat Rev Neurosci. 2018 Jul;19(7):383–4.

10. Cohen SP, Mao J. Neuropathic pain: mechanisms and their clinical implications. BMJ. 2014 Feb 5;348(feb05 6):f7656–f7656.

11. Minikel EV, Painter JL, Dong CC, Nelson MR. Refining the impact of genetic evidence on clinical success. Nature. 2024 May 16;629(8012):624–9.

12. Tam V, Patel N, Turcotte M, Bossé Y, Paré G, Meyre D. Benefits and limitations of genome-wide association studies. Nat Rev Genet. 2019 Aug;20(8):467–84.

13. Johnston KJA, Adams MJ, Nicholl BI, Ward J, Strawbridge RJ, Ferguson A, et al. Genome-wide association study of multisite chronic pain in UK Biobank. Zhu X, editor. PLoS Genet. 2019 Jun 13;15(6):e1008164.

14. Johnston KJA, Ward J, Ray PR, Adams MJ, McIntosh AM, Smith BH, et al. Sex-stratified genome-wide association study of multisite chronic pain in UK Biobank. PLoS Genet. 2021 Apr;17(4):e1009428.

15. Toikumo S, Vickers-Smith R, Jinwala Z, Xu H, Saini D, Hartwell EE, et al. A multi-ancestry genetic study of pain intensity in 598,339 veterans. Nat Med. 2024 Apr;30(4):1075–84.

16. Momi SK, Fabiane SM, Lachance G, Livshits G, Williams FMK. Neuropathic pain as part of chronic widespread pain: environmental and genetic influences. Pain. 2015 Oct;156(10):2100–6.

17. Veluchamy A, Hébert HL, van Zuydam NR, Pearson ER, Campbell A, Hayward C, et al. Association of Genetic Variant at Chromosome 12q23.1 With Neuropathic Pain Susceptibility. JAMA Netw Open. 2021 Dec 1;4(12):e2136560.

18. Packer RJ, Williams AT, Hennah W, Eisenberg MT, Shrine N, Fawcett KA, et al. DeepPheWAS: an R package for phenotype generation and association analysis for phenome-wide association studies. Bioinformatics. 2023 Apr 3;39(4):btad073.

19. NHS Read Browser - TRUD [Internet]. [cited 2024 Sep 5]. Available from: https://isd.digital.nhs.uk/trud/users/guest/filters/0/categories/9/items/8/releases

20. Neuropathic pain | Treatment summaries | BNF content published by NICE [Internet]. [cited 2024 Sep 5]. Available from: https://bnf.nice.org.uk/treatment-summaries/neuropathic-pain/

21. Bycroft C, Freeman C, Petkova D, Band G, Elliott LT, Sharp K, et al. The UK Biobank resource with deep phenotyping and genomic data. Nature. 2018 Oct;562(7726):203–9.

22. Shrine N, Guyatt AL, Erzurumluoglu AM, Jackson VE, Hobbs BD, Melbourne CA, et al. New genetic signals for lung function highlight pathways and chronic obstructive pulmonary disease associations across multiple ancestries. Nat Genet. 2019 Mar;51(3):481–93.

23. Zheng X, Davis JW. SAIGEgds—an efficient statistical tool for large-scale PheWAS with mixed models. Schwartz R, editor. Bioinformatics. 2021 May 5;37(5):728–30.

24. Schizophrenia Working Group of the Psychiatric Genomics Consortium, Bulik-Sullivan BK, Loh PR, Finucane HK, Ripke S, Yang J, et al. LD Score regression distinguishes confounding from polygenicity in genome-wide association studies. Nat Genet. 2015 Mar;47(3):291–5.

25. Yang J, Lee SH, Goddard ME, Visscher PM. GCTA: A Tool for Genome-wide Complex Trait Analysis. The American Journal of Human Genetics. 2011 Jan;88(1):76–82.

26. Yang J, Ferreira T, Morris AP, Medland SE, Madden PAF, Heath AC, et al. Conditional and joint multiple-SNP analysis of GWAS summary statistics identifies additional variants influencing complex traits. Nat Genet. 2012 Apr;44(4):369–75.

27. Wang K, Li M, Hakonarson H. ANNOVAR: functional annotation of genetic variants from high-throughput sequencing data. Nucleic Acids Research. 2010 Sep 1;38(16):e164–e164.

28. Wakefield J. A Bayesian measure of the probability of false discovery in genetic epidemiology studies. Am J Hum Genet. 2007 Aug;81(2):208–27.

29. Pruim RJ, Welch RP, Sanna S, Teslovich TM, Chines PS, Gliedt TP, et al. LocusZoom: regional visualization of genome-wide association scan results. Bioinformatics. 2010 Sep 15;26(18):2336– 7.

30. Reyes-Gibby CC, Wang J, Yeung SCJ, Chaftari P, Yu RK, Hanna EY, et al. Genome-wide association study identifies genes associated with neuropathy in patients with head and neck cancer. Sci Rep. 2018 Jun 8;8(1):8789.

31. Weeks EM, Ulirsch JC, Cheng NY, Trippe BL, Fine RS, Miao J, et al. Leveraging polygenic enrichments of gene features to predict genes underlying complex traits and diseases. Nat Genet. 2023 Aug;55(8):1267–76.

32. Online Mendelian Inheritance in Man, OMIM® [Internet]. McKusick-Nathans Institute of Genetic Medicine, Johns Hopkins University (Baltimore, MD); 2024. Available from: https://omim.org/

33. Gene [Internet] [Internet]. Bethesda (MD): National Library of Medicine (US), National Center for Biotechnology Information; [1988]; 2024. Available from: https://www.ncbi.nlm.nih.gov/gene

34. Sollis E, Mosaku A, Abid A, Buniello A, Cerezo M, Gil L, et al. The NHGRI-EBI GWAS Catalog: knowledgebase and deposition resource. Nucleic Acids Research. 2023 Jan 6;51(D1):D977–85.

35. Sjöstedt E, Zhong W, Fagerberg L, Karlsson M, Mitsios N, Adori C, et al. An atlas of the protein-coding genes in the human, pig, and mouse brain. Science. 2020 Mar 6;367(6482):eaay5947.

36. Ghoussaini M, Mountjoy E, Carmona M, Peat G, Schmidt EM, Hercules A, et al. Open Targets Genetics: systematic identification of trait-associated genes using large-scale genetics and functional genomics. Nucleic Acids Res. 2021 Jan 8;49(D1):D1311–20.

37. Kurki MI, Karjalainen J, Palta P, Sipilä TP, Kristiansson K, Donner KM, et al. FinnGen provides genetic insights from a well-phenotyped isolated population. Nature. 2023 Jan 19;613(7944):508–18.

38. Freidin MB, Tsepilov YA, Palmer M, Karssen LC, Suri P, Aulchenko YS, et al. Insight into the genetic architecture of back pain and its risk factors from a study of 509,000 individuals. Pain. 2019 Jun;160(6):1361–73.

39. Rønn LCB, Hartz BP, Bock E. The neural cell adhesion molecule (NCAM) in development and plasticity of the nervous system. Experimental Gerontology. 1998 Nov;33(7–8):853–64.

40. Duncan BW, Murphy KE, Maness PF. Molecular Mechanisms of L1 and NCAM Adhesion Molecules in Synaptic Pruning, Plasticity, and Stabilization. Front Cell Dev Biol. 2021 Jan 28;9:625340.

41. Kim YH, Kim YH, Shin YK, Jo YR, Park DK, Song M, et al. p75 and neural cell adhesion molecule 1 can identify pathologic Schwann cells in peripheral neuropathies. Ann Clin Transl Neurol. 2019 Jul;6(7):1292–301.

42. Jennings MJ, Kagiava A, Vendredy L, Spaulding EL, Stavrou M, Hathazi D, et al. NCAM1 and GDF15 are biomarkers of Charcot-Marie-Tooth disease in patients and mice. Brain. 2022 Nov 21;145(11):3999–4015.

43. Sakai A, Asada M, Seno N, Suzuki H. Involvement of neural cell adhesion molecule signaling in glial cell line-derived neurotrophic factor-induced analgesia in a rat model of neuropathic pain. Pain. 2008 Jul 15;137(2):378–88.

44. Ko HG, Choi JH, Park DI, Kang SJ, Lim CS, Sim SE, et al. Rapid Turnover of Cortical NCAM1 Regulates Synaptic Reorganization after Peripheral Nerve Injury. Cell Reports. 2018 Jan;22(3):748–59.

45. Knierim E, Hirata H, Wolf NI, Morales-Gonzalez S, Schottmann G, Tanaka Y, et al. Mutations in Subunits of the Activating Signal Cointegrator 1 Complex Are Associated with Prenatal Spinal Muscular Atrophy and Congenital Bone Fractures. The American Journal of Human Genetics. 2016 Mar;98(3):473–89.

46. Rosano KK, Wegner DJ, Shinawi M, Baldridge D, Bucelli RC, Dahiya S, et al. Biallelic ASCC1 variants including a novel intronic variant result in expanded phenotypic spectrum of spinal muscular atrophy with congenital bone fractures 2. American J of Med Genetics Pt A. 2021 Jul;185(7):2190–7.

47. Sharova M, Guseva D, Kurenkov A, Novoselova O, Murtazina A, Skoblov M. Congenital myopathy as a new phenotype caused by two undescribed variants in ASCC1 gene. American J of Med Genetics Pt A. 2022 Oct;188(10):3100–5.

48. Mocci E, Ward K, Perry JA, Starkweather A, Stone LS, Schabrun SM, et al. Genome wide association joint analysis reveals 99 risk loci for pain susceptibility and pleiotropic relationships with psychiatric, metabolic, and immunological traits. Cordell HJ, editor. PLoS Genet. 2023 Oct 16;19(10):e1010977.

49. Song YQ, Karasugi T, Cheung KMC, Chiba K, Ho DWH, Miyake A, et al. Lumbar disc degeneration is linked to a carbohydrate sulfotransferase 3 variant. J Clin Invest. 2013 Nov;123(11):4909–17.

50. Bovonratwet P, Kulm S, Kolin DA, Song J, Morse KW, Cunningham ME, et al. Identification of Novel Genetic Markers for the Risk of Spinal Pathologies: A Genome-Wide Association Study of 2 Biobanks. Journal of Bone and Joint Surgery. 2023 Jun 7;105(11):830–8.

51. Boer CG, Hatzikotoulas K, Southam L, Stefánsdóttir L, Zhang Y, Coutinho De Almeida R, et al. Deciphering osteoarthritis genetics across 826,690 individuals from 9 populations. Cell. 2021 Sep;184(18):4784–4818.e17.

52. Bjornsdottir G, Stefansdottir L, Thorleifsson G, Sulem P, Norland K, Ferkingstad E, et al. Rare SLC13A1 variants associate with intervertebral disc disorder highlighting role of sulfate in disc pathology. Nat Commun. 2022 Feb 2;13(1):634.

53. Martel-Pelletier J, Kwan Tat S, Pelletier JP. Effects of chondroitin sulfate in the pathophysiology of the osteoarthritic joint: a narrative review. Osteoarthritis and Cartilage. 2010 Jun;18:S7–11.

54. Siebert JR, Conta Steencken A, Osterhout DJ. Chondroitin Sulfate Proteoglycans in the Nervous System: Inhibitors to Repair. BioMed Research International. 2014;2014:1–15.

55. Thiele H, Sakano M, Kitagawa H, Sugahara K, Rajab A, Höhne W, et al. Loss of chondroitin 6-O - sulfotransferase-1 function results in severe human chondrodysplasia with progressive spinal involvement. Proc Natl Acad Sci USA. 2004 Jul 6;101(27):10155–60.

56. Tuysuz B, Mizumoto S, Sugahara K, Çelebi A, Mundlos S, Turkmen S. Omani-type spondyloepiphyseal dysplasia with cardiac involvement caused by a missense mutation in CHST3. Clinical Genetics. 2009 Apr;75(4):375–83.

57. Unger S, Lausch E, Rossi A, Mégarbané A, Sillence D, Alcausin M, et al. Phenotypic features of carbohydrate sulfotransferase 3 (CHST3) deficiency in 24 patients: Congenital dislocations and vertebral changes as principal diagnostic features. American J of Med Genetics Pt A. 2010 Oct;152A(10):2543–9.

58. International Multiple Sclerosis Genetics Consortium (IMSGC). Analysis of immune-related loci identifies 48 new susceptibility variants for multiple sclerosis. Nat Genet. 2013 Nov;45(11):1353–60.

59. International Multiple Sclerosis Genetics Consortium, Patsopoulos NA, Baranzini SE, Santaniello A, Shoostari P, Cotsapas C, et al. Multiple sclerosis genomic map implicates peripheral immune cells and microglia in susceptibility. Science. 2019 Sep 27;365(6460):eaav7188.

60. De Guimaraes TAC, Robson AG, De Guimaraes IMC, Laich Y, Aychoua N, Wright G, et al. CDH23 - Associated Usher Syndrome: Clinical Features, Retinal Imaging, and Natural History. Invest Ophthalmol Vis Sci. 2024 Jul 17;65(8):27.

61. Fontanillas P, Kless A, 23andMe Research Team, Bothmer J, Tung JY. Genome-wide association study of pain sensitivity assessed by questionnaire and the cold pressor test. Pain. 2022 Sep;163(9):1763–76.

62. Maegawa GHB. Lysosomal Leukodystrophies Lysosomal Storage Diseases Associated With White Matter Abnormalities. J Child Neurol. 2019 May;34(6):339–58.

63. Balint B, Guerreiro R, Carmona S, Dehghani N, Latorre A, Cordivari C, et al. KCNN2 mutation in autosomal-dominant tremulous myoclonus-dystonia. Euro J of Neurology. 2020 Aug;27(8):1471–7.

64. Callizot N, Guénet JL, Baillet C, Warter JM, Poindron P. The frissonnant Mutant Mouse, a Model of Dopamino-Sensitive, Inherited Motor Syndrome. Neurobiology of Disease. 2001 Jun;8(3):447– 58.

65. Szatanik M, Vibert N, Vassias I, Guénet JL, Eugène D, De Waele C, et al. Behavioral effects of a deletion in Kcnn2, the gene encoding the SK2 subunit of small-conductance Ca2+-activated K+ channels. Neurogenetics. 2008 Oct;9(4):237–48.

66. Kuramoto T, Yokoe M, Kunisawa N, Ohashi K, Miyake T, Higuchi Y, et al. Tremor dominant Kyoto (Trdk) rats carry a missense mutation in the gene encoding the SK2 subunit of small-conductance Ca 2+ -activated K + channel. Brain Research. 2017 Dec;1676:38–45.

67. d’Apolito M, Ceccarini C, Savino R, Adipietro I, Di Bari I, Santacroce R, et al. A Novel KCNN2 Variant in a Family with Essential Tremor Plus: Clinical Characteristics and In Silico Analysis. Genes. 2023 Jun 29;14(7):1380.

68. Qiu H, Miraucourt LS, Petitjean H, Xu M, Theriault C, Davidova A, et al. Parvalbumin gates chronic pain through the modulation of firing patterns in inhibitory neurons. Proc Natl Acad Sci USA. 2024 Jul 2;121(27):e2403777121.

69. Heximer SP, Knutsen RH, Sun X, Kaltenbronn KM, Rhee MH, Peng N, et al. Hypertension and prolonged vasoconstrictor signaling in RGS2-deficient mice. J Clin Invest. 2003 Apr;111(8):1259.

70. Doyen PJ, Beckers P, Brook GA, Hermans E. Regulators of G protein signalling as pharmacological targets for the treatment of neuropathic pain. Pharmacological Research. 2020 Oct;160:105148.

71. Gros P, Milder FJ, Janssen BJC. Complement driven by conformational changes. Nat Rev Immunol. 2008 Jan;8(1):48–58.

72. Fritzinger DC, Benjamin DE. The Complement System in Neuropathic and Postoperative Pain. TOPAINJ. 2016 Sep 30;9(1):26–37.

73. Warwick CA, Keyes AL, Woodruff TM, Usachev YM. The complement cascade in the regulation of neuroinflammation, nociceptive sensitization, and pain. Journal of Biological Chemistry. 2021 Sep;297(3):101085.

74. Tang S, Hu W, Zou H, Luo Q, Deng W, Cao S. The complement system: a potential target for the comorbidity of chronic pain and depression. Korean J Pain. 2024 Apr 1;37(2):91–106.

75. Ebanks R. Mouse complement component C4 is devoid of classical pathway C5 convertase subunit activity. Molecular Immunology. 1996 Feb;33(3):297–309.

76. Schalkwijk J, Zweers MC, Steijlen PM, Dean WB, Taylor G, Van Vlijmen IM, et al. A Recessive Form of the Ehlers–Danlos Syndrome Caused by Tenascin-X Deficiency. N Engl J Med. 2001 Oct 18;345(16):1167–75.

77. Burch GH, Gong Y, Liu W, Dettman RW, Curry CJ, Smith L, et al. Tenascin–X deficiency is associated with Ehlers–Danlos syndrome. Nat Genet. 1997 Sep;17(1):104–8.

78. Chopra P, Tinkle B, Hamonet C, Brock I, Gompel A, Bulbena A, et al. Pain management in the Ehlers–Danlos syndromes. American J of Med Genetics Pt C. 2017 Mar;175(1):212–9.

79. Mao JR, Taylor G, Dean WB, Wagner DR, Afzal V, Lotz JC, et al. Tenascin-X deficiency mimics Ehlers-Danlos syndrome in mice through alteration of collagen deposition. Nat Genet. 2002 Apr;30(4):421–5.

80. Kamada H, Emura K, Yamamoto R, Kawahara K, Uto S, Minami T, et al. Hypersensitivity of myelinated A-fibers via toll-like receptor 5 promotes mechanical allodynia in tenascin-X-deficient mice associated with Ehlers–Danlos syndrome. Sci Rep. 2023 Oct 28;13(1):18490.

81. Sun J, Wang X, Song F, Li D, Gao S, Zhang L, et al. Inhibition of Brd4 alleviates osteoarthritis pain via suppression of neuroinflammation and activation of Nrf2-mediated antioxidant signalling. British J Pharmacology. 2023 Dec;180(24):3194–214.

82. Palomés-Borrajo G, Badia J, Navarro X, Penas C. Nerve Excitability and Neuropathic Pain is Reduced by BET Protein Inhibition After Spared Nerve Injury. The Journal of Pain. 2021 Dec;22(12):1617–30.

83. Borgonetti V, Galeotti N. Combined inhibition of histone deacetylases and BET family proteins as epigenetic therapy for nerve injury-induced neuropathic pain. Pharmacological Research. 2021 Mar;165:105431.

84. Borgonetti V, Meacci E, Pierucci F, Romanelli MN, Galeotti N. Dual HDAC/BRD4 Inhibitors Relieves Neuropathic Pain by Attenuating Inflammatory Response in Microglia After Spared Nerve Injury. Neurotherapeutics. 2022 Sep;19(5):1634–48.

85. Nguyen T, Liu XK, Zhang Y, Dong C. BTNL2, a Butyrophilin-Like Molecule That Functions to Inhibit T Cell Activation. The Journal of Immunology. 2006 Jun 15;176(12):7354–60.

86. Nakajima M, Takahashi A, Kou I, Rodriguez-Fontenla C, Gomez-Reino JJ, Furuichi T, et al. New Sequence Variants in HLA Class II/III Region Associated with Susceptibility to Knee Osteoarthritis Identified by Genome-Wide Association Study. Agarwal S, editor. PLoS ONE. 2010 Mar 18;5(3):e9723.

87. Shi X, Tan H, Le X, Xian H, Li X, Huang K, et al. An expression signature model to predict lung adenocarcinoma-specific survival. Cancer Manag Res. 2018;10:3717–32.

88. Mao Z, Nie Y, Jia W, Wang Y, Li J, Zhang T, et al. Revealing Prognostic and Immunotherapy-Sensitive Characteristics of a Novel Cuproptosis-Related LncRNA Model in Hepatocellular Carcinoma Patients by Genomic Analysis. Cancers. 2023 Jan 16;15(2):544.

89. Sheng J, Liu S, Wang Y, Cui R, Zhang X. The Link between Depression and Chronic Pain: Neural Mechanisms in the Brain. Neural Plast. 2017;2017:9724371.

