## Supplementary Methods for "Genome-wide association study of neuropathic pain phenotypes implicates loci involved in neural cell adhesion, channels, collagen matrix formation and immune regulation"

Supplement

### Supplementary Methods

#### Signal selection

To check for additional independent signals within each locus, we used GCTA 1.94.1 using p-value threshold of 5x10^-8^.

#### Variant to gene mapping

##### Expression quantitative trait loci (eQTLs)

We used coloc.abf to perform approximate Bayes factor colocalisation between the pain GWAS credible sets and eQTLs in eQTLgen blood and the following tissues from GTEx v8: tibial nerve, amygdala, anterior cingulate cortex (BA24), caudate (basal ganglia), cerebellar hemisphere, cerebellum, cortex, frontal cortex (BA9), hippocampus, hypothalamus, nucleus accumbens (basal ganglia), putamen (basal ganglia), spinal cord (cervical c-1), and substantia nigra. We used H4 >80% as evidence of colocalisation.

##### Protein quantitative trait loci (pQTLs)

We included three pQTL datasets in our pQTL analyses: deCODE genetics, which has data on 4719 proteins measured by 4907 aptamers in 35,559 Icelandic individuals; the SCALLOP consortium, which includes data on 90 cardiovascular proteins in 30,931 individuals and; UK Biobank, including data on 1500 proteins measured in 48,195 individuals. We used coloc SuSIE (35,36) to perform approximate Bayes factor colocalisation between the pain GWAS credible sets and pQTLs, where SuSIE could not form credible sets, we used coloc.abf. We used H4 >80% as evidence of colocalisation.

##### Polygenic Priority Score (PoPS)

We used a gene prioritisation method, PoPS (30), to predict causal genes for the sentinel variants from the pain GWAS. The complete set of gene features used in these analyses included 57,543 features: 40,546 derived from gene expression data, 8,718 extracted from a protein-protein interaction network and 8,479 based on pathway membership. In this study, we prioritised genes for all autosomal sentinel variants from the pain GWAS within a 500kb (±250kb) window of the sentinel variant. We reported the top prioritised genes in the region. If no gene was prioritised within a 500kb window of the sentinel variant, we reported any top prioritised genes within a 1Mb window.

##### Nearby Mendelian disease genes

We selected rare Mendelian-disease genes from ORPHANET (https://www.orpha.net/) within ±500kb of a sentinel variant from the pain GWAS that were associated with pain-related diseases. We implicated the gene if any of the following strings were included in the disease name or frequently appeared in human phenotype ontology (HPO) terms for that disease: *pain, neuralgia, neuritis, neuropathy, causalgia, neuropathic, radiculitis, radiculopathy, sciatica, fibromyalgia*. We manually checked the diseases and HPO terms identified for relevance.

##### Nearby mouse knockout orthologs with a pain-related phenotype

We identified nearby genes within +/- 500kb of each GWAS sentinel and, using the Mouse Genome Informatics (MGI) database, identified mouse orthologs containing at least one allele classified as either a conditional or knock-out allele conferring a ‘nervous system’ or ‘behaviour/neurological phenotype’. We then manually reviewed the phenotypes, identifying pain and/or neuropathy.

##### Functional annotation of credible set variants

We used Variant Effect Predictor (VEP) to annotate variants in the 99% credible sets from the pain GWAS. We implicated a gene as putatively causal if any credible set variant had >50% posterior probability and if it additionally was a missense variant, annotated as "deleterious" by SIFT, "damaging" by PolyPhen-2 or had a CADD PHRED score ≥20.

##### Rare variant analysis from whole-exome sequencing

Using UK Biobank whole exome sequencing data within +/-500kb of each GWAS sentinel variant, we tested rare single-variant and gene-based associations using the neuropathic pain phenotypes developed for GWAS testing in addition to, back pain (see supplementary text alongside Supplementary Tables 2-6) and multisite chronic pain, defined following (13). The single variant tests were conducted using REGENIE v3.2.5 for variants with MAC ≥3 and for phenotypes with over 4000 cases, under additive, dominant and recessive genetic models, adjusted for age, sex, genotyping array and the first 10 principal components of ancestry.

We used REGENIE v3.3 (37) to perform gene-based testing for all phenotypes with over 1000 cases using two annotation masks (38): M1(predicted loss-of-function [pLoF]) and M3 (pLoF and likely deleterious missense). “Likely deleterious” was defined as predicted deleterious by all five algorithms: SIFT, PolyPhen2 HDIV, PolyPhen2 HVAR, LRT and MutationTaster. We considered up to five minor allele frequency (MAF) thresholds: 1%, 0.1%, 0.01%, 0.001%, singleton. We included variants with at least 90% of the genotypes having a read depth >10 for quality control filtering and excluded variants with a minor allele count (MAC) <3. For the gene-based associating testing, we used the following options: ‘--vc-tests acatv, skato --joint acat’ for binary traits. Age, sex, genotyping array and the first 10 principal components of ancestry were included as covariates. We specified the --rgc-gene-p option to calculate a single p-value per gene using the different combinations of mask, MAF and test. The variant-to-gene mapping single rare variant association criterion was met if the +/-500kb region surrounding the GWAS sentinel contained a rare variant single variant association with P<5x10^-6^ and the variant-to-gene mapping gene-based association criterion was met if the +/-500kb region surrounding the GWAS sentinel contained a gene meeting P<5x10^-4^ for gene-based testing.

### Phenotyping

Pain phenotypes were developed using DeepPheWAS. DeepPheWAS combines lists of related codes together to make new lists. These lists can be lists of cases or controls that can then further be combined to make phenotypes. The most common lists of codes are clinical codes from either ICD10/9, Read V2/V3, self-reported illness or operation or OPSC4. The two other variations are prescription drug lists, coded as key search terms given the limitation in UK Biobank drug recording in the linked primary care records, and field ID lists. Field ID lists describe a field-id within the UK biobank. Alongside the ID, there are individual codes to extract.

Each phenotype used in DeepPheWAS has a map showing how each list is combined. Each phenotype used in this paper was developed using these tools, and the map (formulae) is described below. Help to understand how to read those formulae alongside some special use cases are detailed directly below.

#### How to interpret notation

The names within each formula are lists of related codes **AND**, **OR**, **NOT** are Boolean logic operators, **OR** is represented by | in the formulae. N refers to the number of codes required. Formulae are read and evaluated from left to right. ΔT refers to the time gap in days between the left and right object’s first recorded code and acts as a filter. A positive time gap would indicate that the right object occurred after the left, so in the example below, a case must have 1 code for anti_epileptic_pain_drugs and NOT any code for epilepsy that occurs less than 365 days after the first code for anti_epileptic_pain_drugs.

$$case_{1}\boldsymbol{=}anti\_epileptic\_pain\_drugs\_N1\boldsymbol{NOT (}epilepsy\_N1 \Delta T<365 )$$

The time gap can also be a range, in the example below, the object on the right must have occurred from 30 days before to less than 365 days after the object on the left.

$$case_{1}\boldsymbol{=}anti\_epileptic\_pain\_drugs\_N1\boldsymbol{NOT (}epilepsy\_N1 >-30 \Delta T<365 )$$

Most use of the time gaps are not this complicated and should be relatively easy to understand.

#### Control populations

Phenotypes that were used only to define other phenotypes and were not analysed separately do not have controls. Where there are controls the control population is either nested within a phenotype or is population level. Population level controls are called primary_care_pop in the formulae which consists of everyone with available primary care data. Exclusions are then made to either the nested or population controls to get the final controls for each phenotype.

#### Collapsed phenotypes

The collapsed phenotypes all-cause, central, and peripheral neuropathic pain are not included here as they have been described previously and do not lend themselves to the formulae notation, in all three cases primary_care_pop is used as their starting control population.

#### Drugs for neuropathic pain

Prescriptions for medication that is used to treat neuropathic pain is used for many of the neuropathic pain phenotypes these are not analysed separately and so have no controls. There are two used for defining neuropathic pain phenotypes, Neuropathic_pain_drugs and Neuropathic_pain_drugs_wider.

#### Composite concepts formula

##### Neuropathic_pain_drugs

$$case_{1}\boldsymbol{=}anti\_epileptic\_pain\_drugs\_N1\boldsymbol{NOT (}epilepsy\_N1 \Delta T<365)$$

$$case_{2}\boldsymbol{=}anti\_depressant\_pain\_drugs\_N1$$

$case_{3}\boldsymbol{=}other\_neuropathic\_pain\_drugs\_N1$

##### Neuropathic_pain_drugs_wider

$case_{1}\boldsymbol{=}anti\_epileptic\_pain\_drugs\_N1\boldsymbol{NOT (}epilepsy\_N1 \Delta T<365)$

$$case_{2}\boldsymbol{=}anti\_depressant\_pain\_drugs\_N1$$

$case_{3}\boldsymbol{=}other\_neuropathic\_pain\_drugs\_N1$

$$case_{4}\boldsymbol{=}other\_pain\_drugs\_N1$$

##### Carpel_tunnel_wider

$$case_{1}\mathbf{=}carpal\_tunnel\_syndrome\_N1|carpal\_tunnel\_surgery\_N1|ever\_had\_carpal\_tunnel\_syndrome\_N1$$

##### Spinal_impingement_wider

$$case_{1}\boldsymbol{=}spinal\_impingement\_conditions\_N1|spinal\_operations\_OPCS\_SR\_N1$$

##### Parkinsons_cases

$$case_{1}\boldsymbol{=parkinsons}\_N1\boldsymbol{AND (parkinsons\_drugs}\_N1 \Delta T>-30) NOT secondary\_parkinsons\_N1$$

#### Phenotype Formula

##### P2200.13 Alzheimers with neuropathic pain

$case_{1}\boldsymbol{=}Alzheimers\_disease\_N1\boldsymbol{AND (}central\_neuropathic\_pain\_N1 | neuropathic\_pain\_drugs\_N2 \Delta T>0 )$

$$control\boldsymbol{=}Alzheimers\_disease\_N1\boldsymbol{NOT (}central\_neuropathic\_pain\_N1 | neuropathic\_pain\_drugs\_wider N1|other\_dementia\_N1 )$$

##### P2228 Back pain

$case_{1}\boldsymbol{=}back\_pain\_N2$

$$control\boldsymbol{=}primary\_care\_pop\_N1 \boldsymbol{NOT}back\_pain\_N1$$

##### P2200.42 Complex regional pain syndrome

$$case_{1}\boldsymbol{=(}complex\_regional\_pain\_syndrome\_N1 \boldsymbol{|}Ever\_had\_complex\_regional\_pain\_syndrome\_N1\boldsymbol{)}$$

$$control\boldsymbol{=}primary\_care\_pop\_N1\boldsymbol{NOT(}complex\_regional\_pain\_syndrome\_N1 \boldsymbol{|} Ever\_had\_complex\_regional\_pain\_syndrome\_N1\boldsymbol{)}$$

##### P2200.44 Fabry’s disease with neuropathic pain

$case_{1}\boldsymbol{=}fabry\_N1\boldsymbol{AND (}peripheral\_neuropathic\_pain\_N1 | neuropathic\_pain\_drugs\_N2 \Delta T>0)$

$$control\boldsymbol{=}fabry\_N1\boldsymbol{NOT (}peripheral\_neuropathic\_pain\_N1 | neuropathic\_pain\_drugs\_wider\_N1)$$

##### P2200.39 Clinical fibromyalgia

$$case_{1}\boldsymbol{=(}fibromyalgia\_N2 \boldsymbol{|}Ever\_had\_fibromyalgia\_syndrome\_N1\boldsymbol{)}$$

$$control\boldsymbol{=}primary\_care\_pop\_N1\boldsymbol{NOT(}fibromyalgia\_N1 \boldsymbol{|}Ever\_had\_fibromyalgia\_syndrome\_N1\boldsymbol{)}$$

##### P2200.391 Fibromyalgia pain clinical (+drugs)

$case_{1}\boldsymbol{=(}fibromyalgia\_N2 \boldsymbol{|}Ever\_had\_fibromyalgia\_syndrome\_N1\boldsymbol{) AND (}peripheral\_neuropathic\_pain\_N1 | neuropathic\_pain\_drugs\_N2 \Delta T>-60)$

$$control\boldsymbol{=(}fibromyalgia\_N1 \boldsymbol{|}Ever\_had\_fibromyalgia\_syndrome\_N1\boldsymbol{) NOT (}peripheral\_neuropathic\_pain\_N1 | neuropathic\_pain\_drugs\_wider\_N1)$$

##### P2200.37 Guillain-Barre syndrome with neuropathic pain

$case_{1}\boldsymbol{=}guillainBarre\_N1\boldsymbol{AND (}peripheral\_neuropathic\_pain\_N1 | neuropathic\_pain\_drugs\_N2 \Delta T>0)$

$$control\boldsymbol{=}guillainBarre\_N1\boldsymbol{NOT (}peripheral\_neuropathic\_pain\_N1 | neuropathic\_pain\_drugs\_wider\_N1)$$

##### P2200.12 Huntington's with neuropathic pain

$case_{1}\boldsymbol{=}huntingtons\_N1\boldsymbol{AND (}central\_neuropathic\_pain\_N1 | neuropathic\_pain\_drugs\_N2 \Delta T>0)$

$$control\boldsymbol{=}huntingtons\_N1\boldsymbol{NOT (}central\_neuropathic\_pain\_N1 | neuropathic\_pain\_drugs\_wider\_N1)$$

##### P2200.36 Leprosy with neuropathic pain

$case_{1}\boldsymbol{=(}leprosy\_N1 \boldsymbol{|}leprosy\_neuropathy\boldsymbol{\_}N1\boldsymbol{) AND (}peripheral\_neuropathic\_pain\_N1 | neuropathic\_pain\_drugs\_N2 \Delta T>-60)$

$$control\boldsymbol{=}leprosy\_N1\boldsymbol{NOT (}peripheral\_neuropathic\_pain\_N1 | neuropathic\_pain\_drugs\_wider\_N1)$$

##### P2200.15 Multiple sclerosis with neuropathic pain

$case_{1}\boldsymbol{=}multiple\_sclerosis\_N1\boldsymbol{AND (}central\_neuropathic\_pain\_N1 | neuropathic\_pain\_drugs\_N2 \Delta T>-60)$

$$control\boldsymbol{=}multiple\_sclerosis\_N1\boldsymbol{NOT (}central\_neuropathic\_pain\_N1 | neuropathic\_pain\_drugs\_wider\_N1)$$

##### P2200.43 Neurofibromatosis type 2 schwannomatosis schwannomas with neuropathic pain

$case_{1}\boldsymbol{=}neurofibromatosis\_type2\_N1\boldsymbol{AND (}peripheral\_neuropathic\_pain\_N1 | neuropathic\_pain\_drugs\_N2 \Delta T>-30)$

$$control\boldsymbol{=}neurofibromatosis\_type2\_N1\boldsymbol{NOT (}peripheral\_neuropathic\_pain\_N1 | neuropathic\_pain\_drugs\_wider\_N1)$$

##### P2200.20 Neuromyelitis Optica with neuropathic pain

$case_{1}\boldsymbol{=}neuromyelitis\_optica\_N1\boldsymbol{AND (}central\_neuropathic\_pain\_N1 | neuropathic\_pain\_drugs\_N2 \Delta T>-30)$

$$control\boldsymbol{=}neuromyelitis\_optica\_N1\boldsymbol{NOT (}central\_neuropathic\_pain\_N1 | neuropathic\_pain\_drugs\_wider\_N1)$$

##### P2200.41 Neuropathic back pain

$case_{1}\boldsymbol{=}back\_pain\_N2\boldsymbol{AND (}peripheral\_neuropathic\_pain\_N1 | neuropathic\_pain\_drugs\_N2 \Delta T>0)$

$$control\boldsymbol{=}back\_pain\_N1\boldsymbol{NOT (}peripheral\_neuropathic\_pain\_N1 | neuropathic\_pain\_drugs\_wider\_N1)$$

##### P2200.35 Neuropathic pain due to HIV infection

$case_{1}\boldsymbol{=}HIV\_N2\boldsymbol{AND (}peripheral\_neuropathic\_pain\_N1 | neuropathic\_pain\_drugs\_N2 \Delta T>0)$

$case_{2}\boldsymbol{=}HIV\_neuropathy\_N1\boldsymbol{AND (}peripheral\_neuropathic\_pain\_N1 | neuropathic\_pain\_drugs\_N2 \Delta T>-30)$

$$control\boldsymbol{=}HIV\_N1\boldsymbol{NOT (}peripheral\_neuropathic\_pain\_N1 | neuropathic\_pain\_drugs\_wider\_N1 | HIV\_neuropathy\_N1)$$

##### P2200.53 Other neuropathy and compression pain

$case_{1}\boldsymbol{=}other\_neuropathy\_and\_compression\_N1\boldsymbol{AND (}peripheral\_neuropathic\_pain\_N1 | neuropathic\_pain\_drugs\_N2 \Delta T>-30)$

$$control\boldsymbol{=}other\_neuropathy\_and\_compression\_N1\boldsymbol{NOT (}peripheral\_neuropathic\_pain\_N1 | neuropathic\_pain\_drugs\_wider\_N1)$$

##### P2200.46 Painful carpal tunnel syndrome

$case_{1}\boldsymbol{=(}carpal\_tunnel\_syndrome\_N2 \boldsymbol{|} Ever\_had\_carpal\_tunnel\_syndrome\_N1\boldsymbol{) AND (}peripheral\_neuropathic\_pain\_N1 | neuropathic\_pain\_drugs\_N2 \Delta T>-60)$

$$control\boldsymbol{=}carpal\_tunnel\_wider\_N1\boldsymbol{NOT (}peripheral\_neuropathic\_pain\_N1 | neuropathic\_pain\_drugs\_wider\_N1)$$

##### P2200.31 Painful diabetic neuropathy

$case_{1}\boldsymbol{=}diabetes\_all\_N3 \boldsymbol{AND (}peripheral\_neuropathic\_pain\_N1 | neuropathic\_pain\_drugs\_N2 \Delta T>1)$

$case_{2}\boldsymbol{=}diabetic\_neuropathy\_generic\_N1 \boldsymbol{AND (}peripheral\_neuropathic\_pain\_N1 | neuropathic\_pain\_drugs\_N2 | other\_pain\_drugs\_N2 \Delta T>-30)$

$case_{3}\boldsymbol{=}diabetic\_neuropathy\_N1$

$$control\boldsymbol{=}diabetes\_all\_N3 \boldsymbol{AND (}peripheral\_neuropathic\_pain\_N1 | neuropathic\_pain\_drugs\_wider\_N1 | other\_pain\_drugs\_N1 | diabetic\_neuropathy\_generic\_N1 |$$

$Ever\_had\_diabetic\_neuropathy\_N1)$

##### P2200.48 Painful discopathy

$case_{1}\boldsymbol{=}discopathy\_N1\boldsymbol{AND (}peripheral\_neuropathic\_pain\_N1 | neuropathic\_pain\_drugs\_N2 \Delta T>0)$

$$control\boldsymbol{=}discopathy\_N1\boldsymbol{NOT (}peripheral\_neuropathic\_pain\_N1 | neuropathic\_pain\_drugs\_wider\_N1)$$

##### P2200.45 Painful peripheral nerve injury or entrapment

$case_{1}\boldsymbol{=}peripheral\_nerve\_injury\_entrapment\_N1\boldsymbol{AND (}peripheral\_neuropathic\_pain\_N1 | neuropathic\_pain\_drugs\_N2 \Delta T>-60)$

$$control\boldsymbol{=}peripheral\_nerve\_injury\_entrapment\_N1\boldsymbol{NOT (}peripheral\_neuropathic\_pain\_N1 | neuropathic\_pain\_drugs\_wider\_N1)$$

##### P2200.47 Painful spinal nerve impingement

$case_{1}\boldsymbol{=}spinal\_impingement\_conditions\_N1\boldsymbol{AND (}peripheral\_neuropathic\_pain\_N1 | neuropathic\_pain\_drugs\_N2 \Delta T>-60)$

$$control\boldsymbol{=}spinal\_impingement\_conditions\_N1\boldsymbol{NOT (}peripheral\_neuropathic\_pain\_N1 | neuropathic\_pain\_drugs\_wider\_N1|spinal\_nerve\_impingement\_wider\_N1)$$

##### P2200.49 Painful spinal stenosis

$case_{1}\boldsymbol{=}spinal\_stenosis\_N1\boldsymbol{AND (}peripheral\_neuropathic\_pain\_N1 | neuropathic\_pain\_drugs\_N2 \Delta T>-60)$

$$control\boldsymbol{=}spinal\_stenosis\_N1\boldsymbol{NOT (}peripheral\_neuropathic\_pain\_N1 | neuropathic\_pain\_drugs\_wider\_N1)$$

##### P2200.50 Painful spondylosis

$case_{1}\boldsymbol{=}spondylosis\_N1\boldsymbol{AND (}peripheral\_neuropathic\_pain\_N1 | neuropathic\_pain\_drugs\_N2 \Delta T>-60)$

$$control\boldsymbol{=}spondylosis\_N1\boldsymbol{NOT (}peripheral\_neuropathic\_pain\_N1 | neuropathic\_pain\_drugs\_wider\_N1)$$

##### P2200.11 Parkinson's (simple) with neuropathic pain

$case_{1}\boldsymbol{=}parkinsons\_N1\boldsymbol{AND (}central\_neuropathic\_pain\_N1 | neuropathic\_pain\_drugs\_N2 \Delta T>0)$

$$control\boldsymbol{=}parkinsons\_N1\boldsymbol{NOT (}central\_neuropathic\_pain\_N1 | neuropathic\_pain\_drugs\_wider\_N1)$$

##### P2200.111 Parkinson's with neuropathic pain

$case_{1}\boldsymbol{=}parkinsons\_cases\_N1\boldsymbol{AND (}central\_neuropathic\_pain\_N1 | neuropathic\_pain\_drugs\_N2 \Delta T>0)$

$$control\boldsymbol{=}parkinsons\_cases\_N1\boldsymbol{NOT (}central\_neuropathic\_pain\_N1 | neuropathic\_pain\_drugs\_wider\_N1)$$

##### P2200.34 Post-amputation neuropathic pain

$case_{1}\boldsymbol{=}amputation\_N1\boldsymbol{AND (}peripheral\_neuropathic\_pain\_N1 | neuropathic\_pain\_drugs\_N2 \Delta T>0)$

$case_{2}\boldsymbol{=}phantom\_limb\_\_N1$

$$control\boldsymbol{=}amputation\_N1\boldsymbol{NOT (}peripheral\_neuropathic\_pain\_N1 | neuropathic\_pain\_drugs\_wider\_N1 | phantom\_limb\_\_N1)$$

##### P2200.33 Post-herpetic neuralgia

$case_{1}\boldsymbol{=}herpes\_N1\boldsymbol{AND (}peripheral\_neuropathic\_pain\_N1 | neuropathic\_pain\_drugs\_N2 \Delta T>0)$

$case_{2}\boldsymbol{=(}post\_herpetic\_neuralgia\_\_N1 | Ever\_had\_post\_herpetic\_neuralgia\_N1)$

$$control\boldsymbol{=}herpes\_N1\boldsymbol{NOT (}peripheral\_neuropathic\_pain\_N1 | neuropathic\_pain\_drugs\_wider\_N1 | post\_herpetic\_neuralgia\_\_N1 | Ever\_had\_post\_herpetic\_neuralgia\_N1 )$$

##### P2200.40 Post-surgical neuropathic pain

$case_{1}\boldsymbol{=(}post\_surgical\_neuropathic\_pain\_N1 \boldsymbol{|} Ever had chronic post surgical pain\_N1\boldsymbol{) AND (}peripheral\_neuropathic\_pain\_N1 | neuropathic\_pain\_drugs\_N2 \Delta T>0)$

$$control\boldsymbol{=(}post\_surgical\_neuropathic\_pain\_N1 \boldsymbol{|} Ever had chronic post surgical pain\_N1\boldsymbol{) NOT (}peripheral\_neuropathic\_pain\_N1 | neuropathic\_pain\_drugs\_wider\_N1)$$

##### P2200.51 Sciatica clinical

$$case_{1}\boldsymbol{=}sciatica\_N2$$

$$control\boldsymbol{=}primary\_care\_pop\_N1\boldsymbol{NOT}sciatica\boldsymbol{\_}N1$$

##### P2200.511 Sciatica clinical + drugs

$case_{1}\boldsymbol{=}sciatica\_N1\boldsymbol{AND (}peripheral\_neuropathic\_pain\_N1 | neuropathic\_pain\_drugs\_N2 \Delta T>-60)$

$$control\boldsymbol{=}sciatica\_N1\boldsymbol{NOT (}peripheral\_neuropathic\_pain\_N1 | neuropathic\_pain\_drugs\_wider\_N1)$$

##### P2200.16 Spinal cord injury associated neuropathic pain

$case_{1}\boldsymbol{=}spinal\_cord\_injury\_N1\boldsymbol{AND (}central\_neuropathic\_pain\_N1 | neuropathic\_pain\_drugs\_N2 \Delta T>0)$

$$control\boldsymbol{=}spinal\_cord\_injury\_N1\boldsymbol{NOT (}central\_neuropathic\_pain\_N1 | neuropathic\_pain\_drugs\_wider\_N1)$$

##### P2200.14 Stroke associated neuropathic pain

$case_{1}\boldsymbol{=}stroke\_N1\boldsymbol{AND (}central\_neuropathic\_pain\_N1 | neuropathic\_pain\_drugs\_N2 \Delta T>0)$

$$control\boldsymbol{=}stroke\_N1\boldsymbol{NOT (}central\_neuropathic\_pain\_N1 | neuropathic\_pain\_drugs\_wider\_N1 | stroke\_pain\_N1)$$

##### P2200.18 Syringomyelia with neuropathic pain

$case_{1}\boldsymbol{=}syringomyelia\_N1\boldsymbol{AND (}central\_neuropathic\_pain\_N1 | neuropathic\_pain\_drugs\_N2 \Delta T>-60)$

$$control\boldsymbol{=}syringomyelia\_N1\boldsymbol{NOT (}central\_neuropathic\_pain\_N1 | neuropathic\_pain\_drugs\_wider\_N1)$$

##### P2200.19 Transverse myelitis with neuropathic pain

$case_{1}\boldsymbol{=}transverse\_myelitis\_N1\boldsymbol{AND (}central\_neuropathic\_pain\_N1 | neuropathic\_pain\_drugs\_N2 \Delta T>-30)$

$$control\boldsymbol{=}transverse\_myelitis\_N1\boldsymbol{NOT (}central\_neuropathic\_pain\_N1 | neuropathic\_pain\_drugs\_wider\_N1)$$

##### P2200.17 Traumatic brain injury associated neuropathic pain

$case_{1}\boldsymbol{=}brain\_injury\_N1\boldsymbol{AND (}central\_neuropathic\_pain\_N1 | neuropathic\_pain\_drugs\_N2 \Delta T>0)$

$$control\boldsymbol{=}brain\_injury\_N1\boldsymbol{NOT (}central\_neuropathic\_pain\_N1 | neuropathic\_pain\_drugs\_wider\_N1 | stroke\_pain\_N1)$$

##### P2200.32 Trigeminal neuralgia

$$case_{1}\boldsymbol{=}trigeminal\_neuralgia\_N1$$

$$control\boldsymbol{=}primary\_care\_pop\_N1\boldsymbol{NOT}trigeminal\_neuralgia\boldsymbol{\_}N1$$

### Supplementary Figures

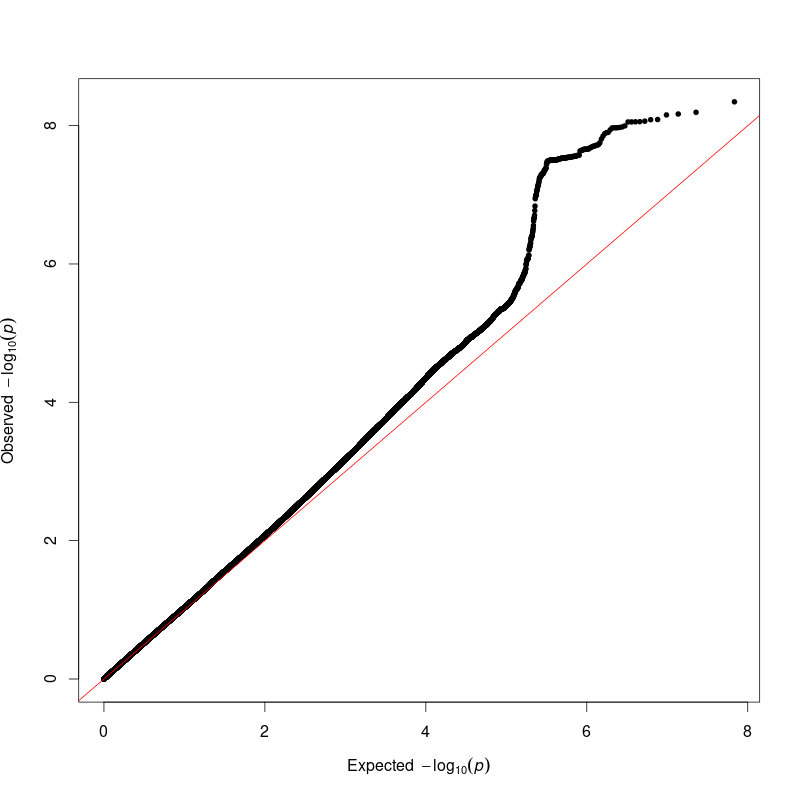

Supplementary Figure 1: QQ plot of GWAS results for P2200_EUR (All-cause neuropathic Pain), LD intercept 1.02

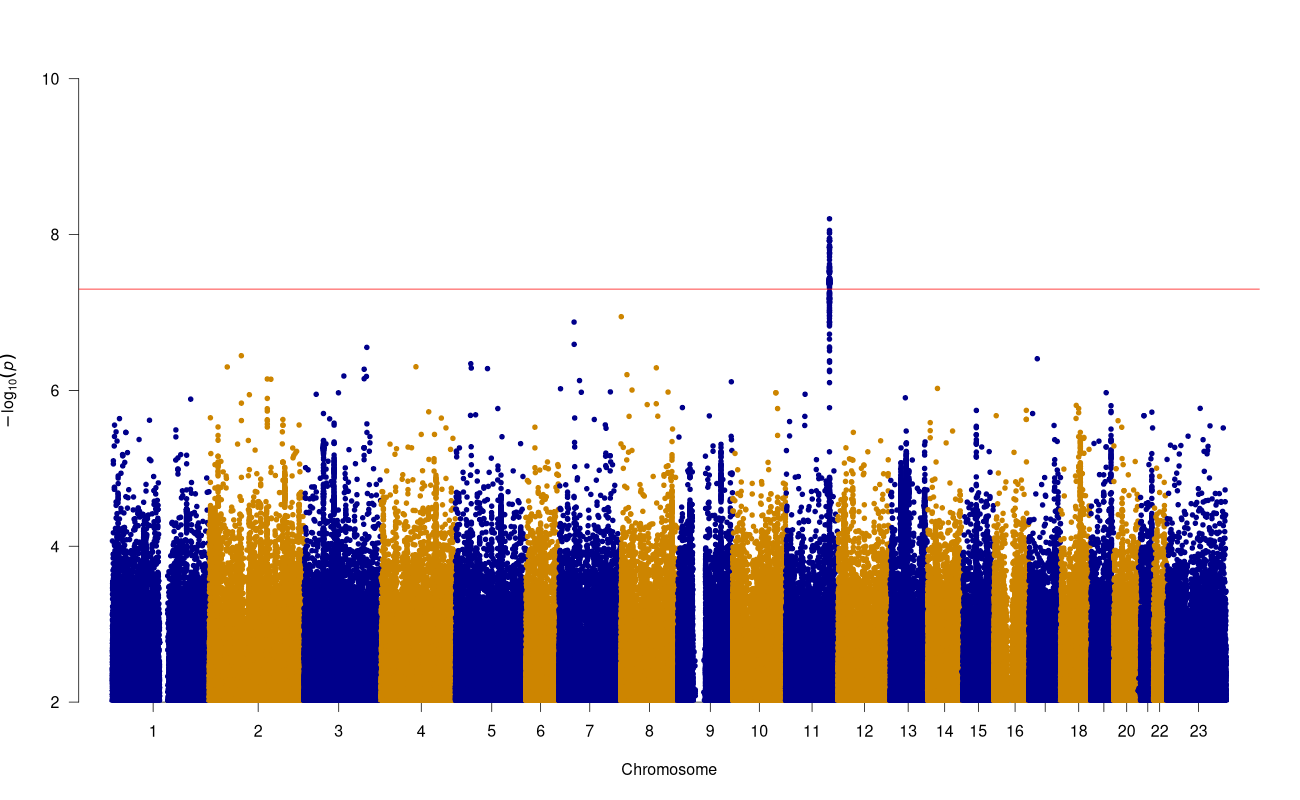

Supplementary Figure 2: Manhattan plot of GWAS results for P2200_EUR (All-cause neuropathic Pain) (red line=5x10^-8^, blue line=5x10^-7^)

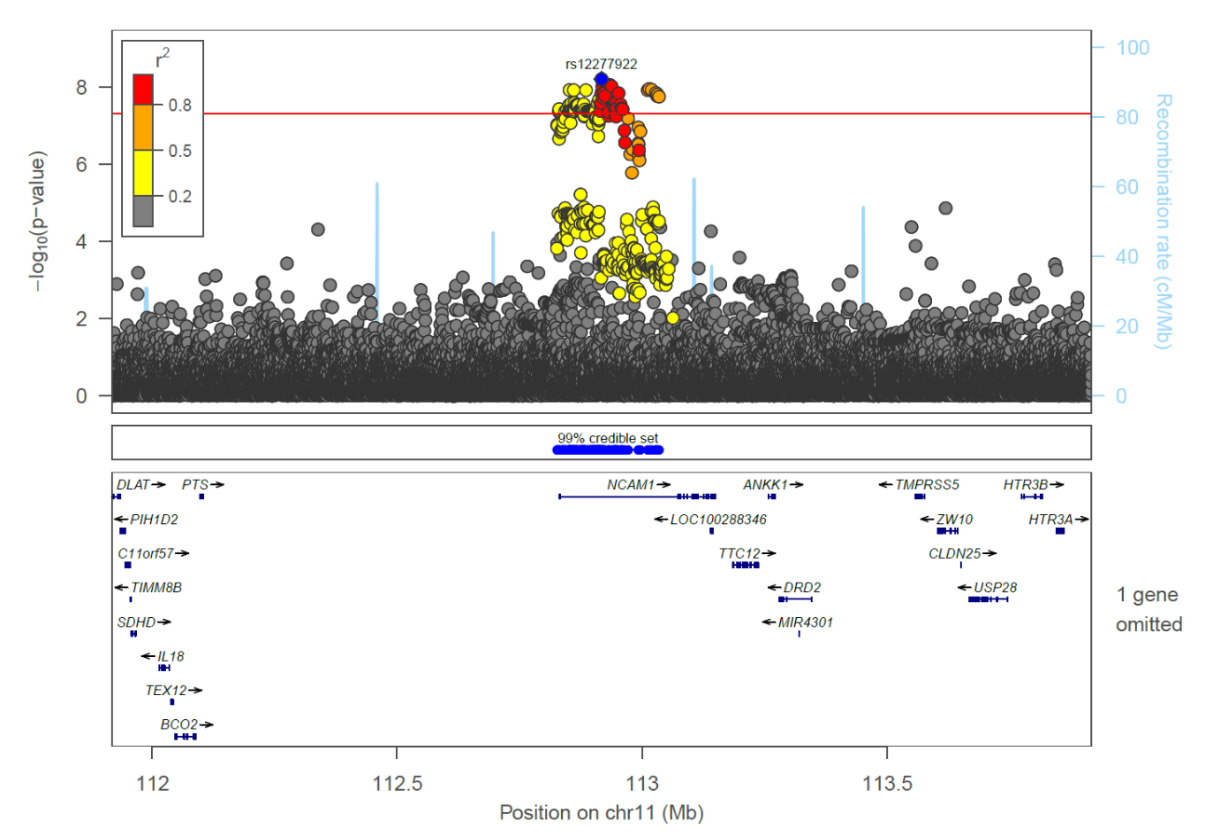

Supplementary Figure 3**:** Region plot for P2200_EUR all-cause neuropathic pain *NCAM1* locus, sentinel SNP labelled with a blue diamond, credible set position shown by blue dots below the region plot.

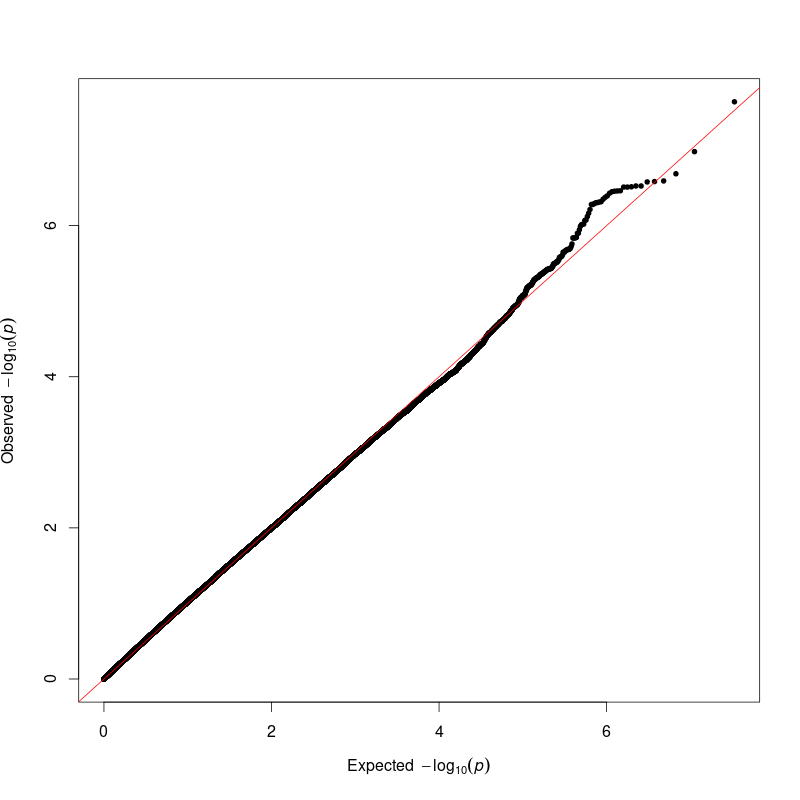

Supplementary Figure 4: QQ plot of GWAS results for P2200_AFR (All-cause neuropathic Pain), LD intercept 1.02

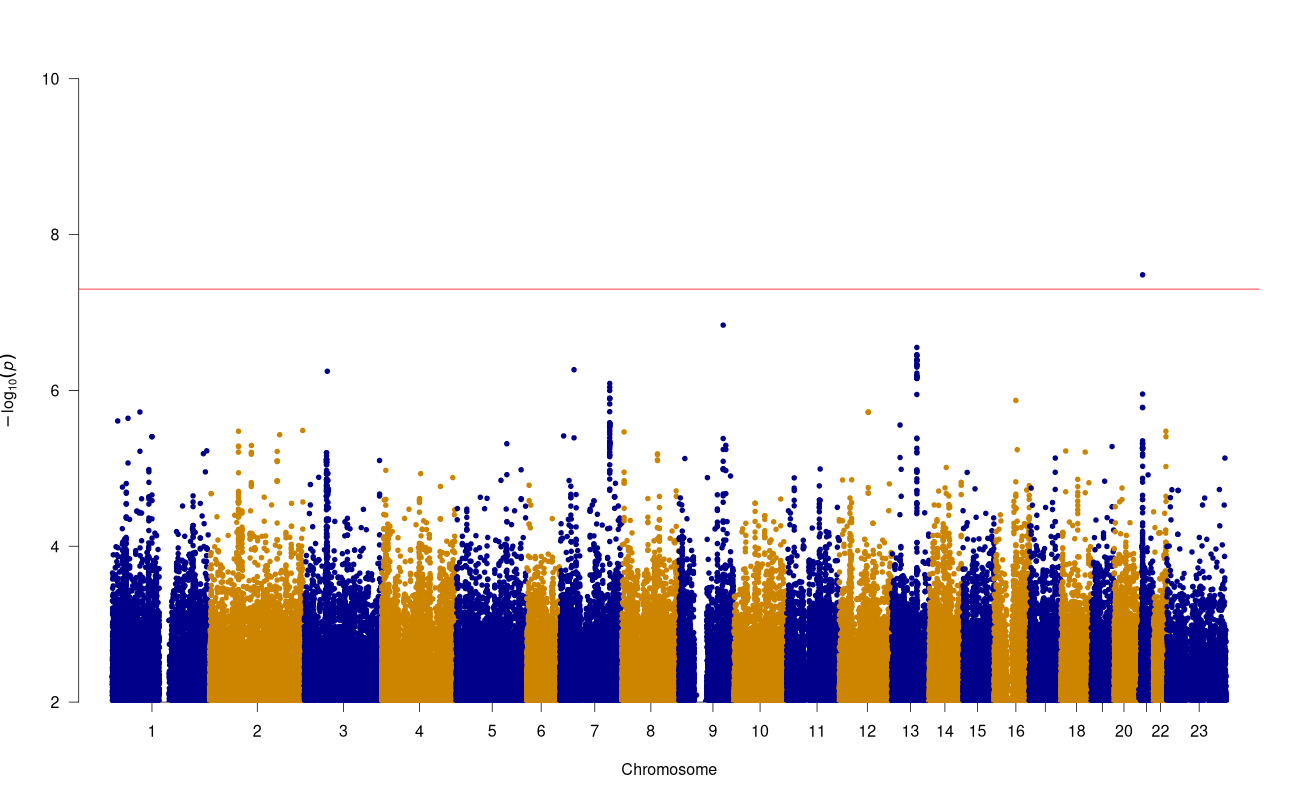
Supplementary Figure 5: Manhattan plot of GWAS results for P2200_AFR (All-cause neuropathic Pain) (red line=5x10^-8^, blue line=5x10^-7^)

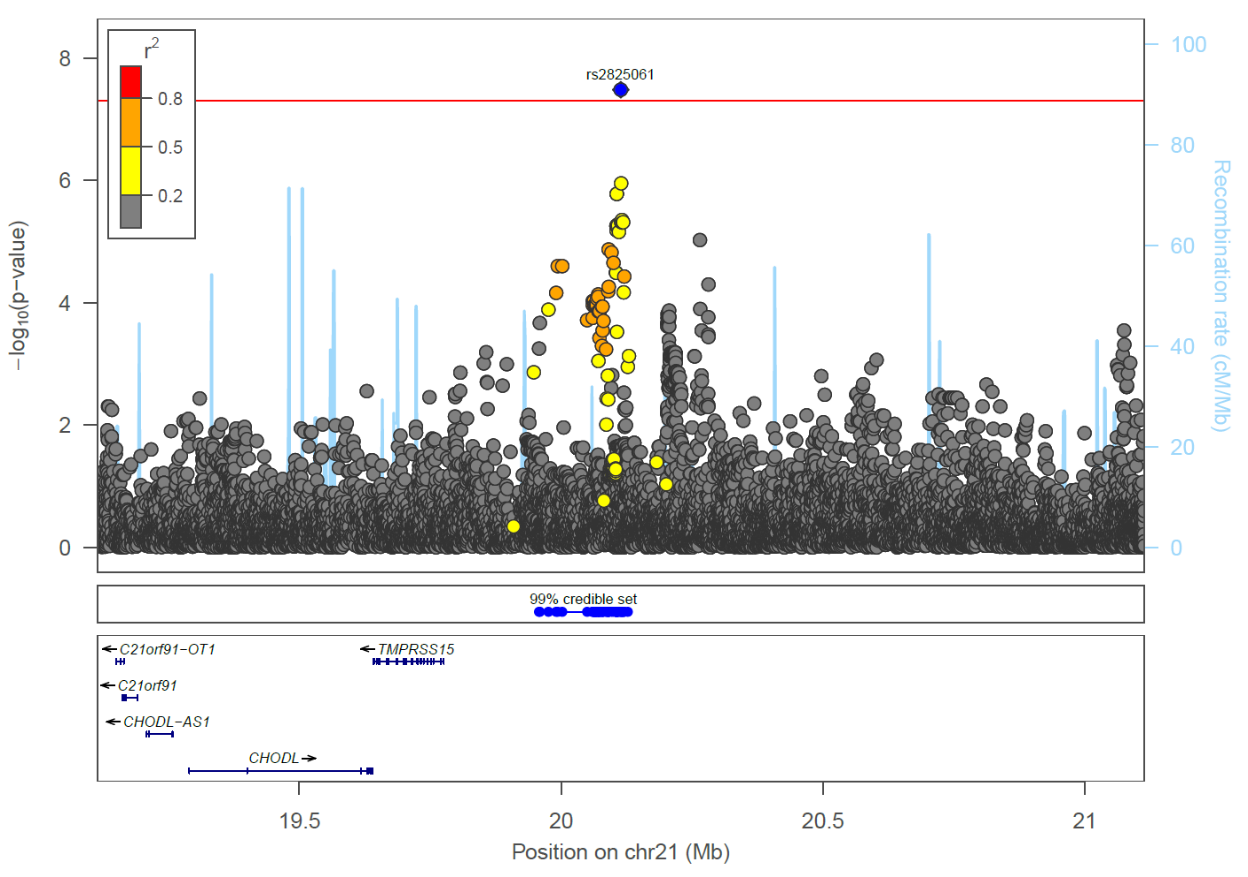

Supplementary Figure 6**:** Region plot for P2200_AFR all-cause neuropathic pain *MIR548XHG* locus, sentinel SNP labelled with a blue diamond, credible set position shown by blue dots below the region plot.

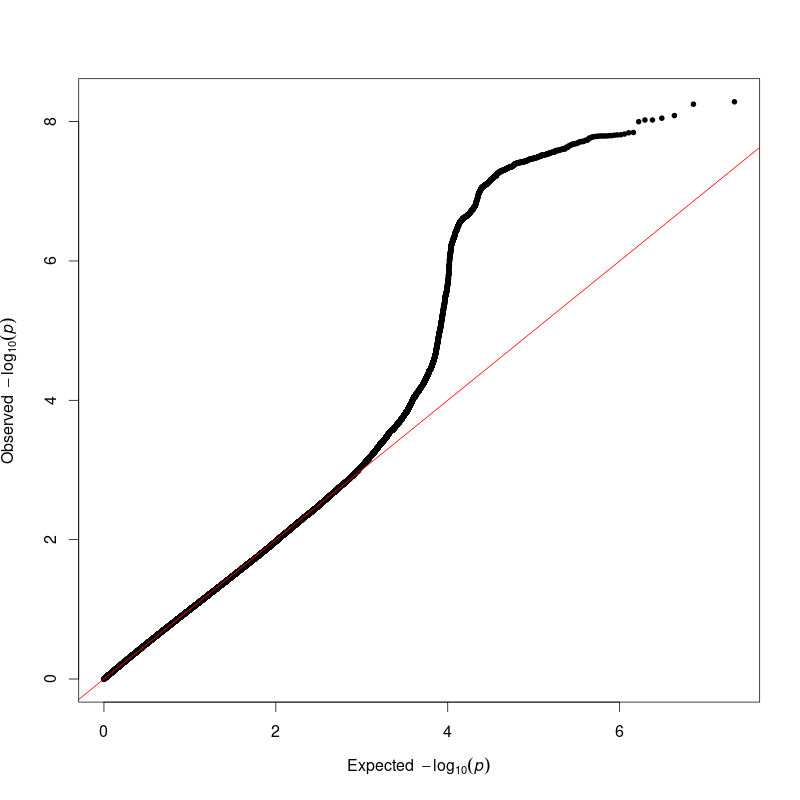

Supplementary Figure 7: QQ plot of GWAS results for P2200.1_EUR (Central neuropathic pain), LD intercept 1.01

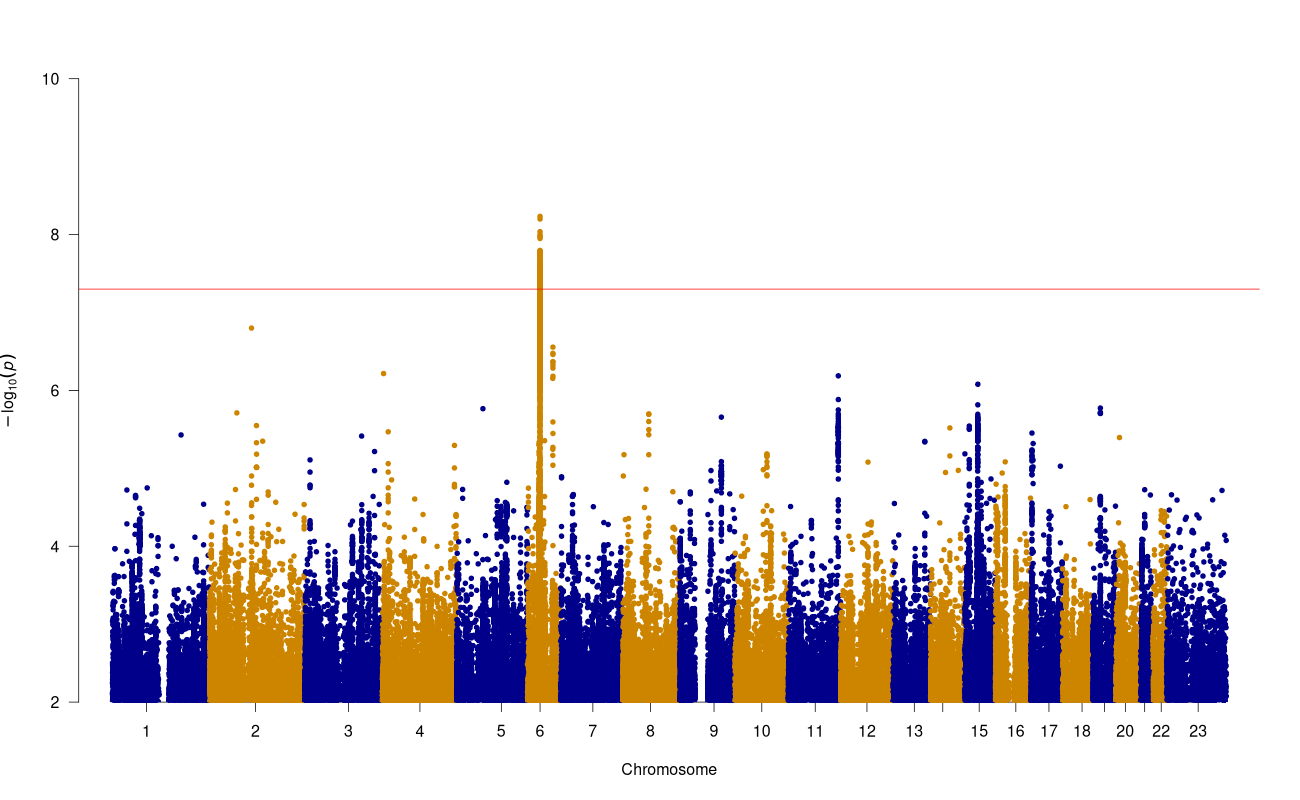

Supplementary Figure 8: Manhattan plot of GWAS results for P2200.1_EUR (Central neuropathic pain) (red line=5x10^-8^, blue line=5x10^-7^)

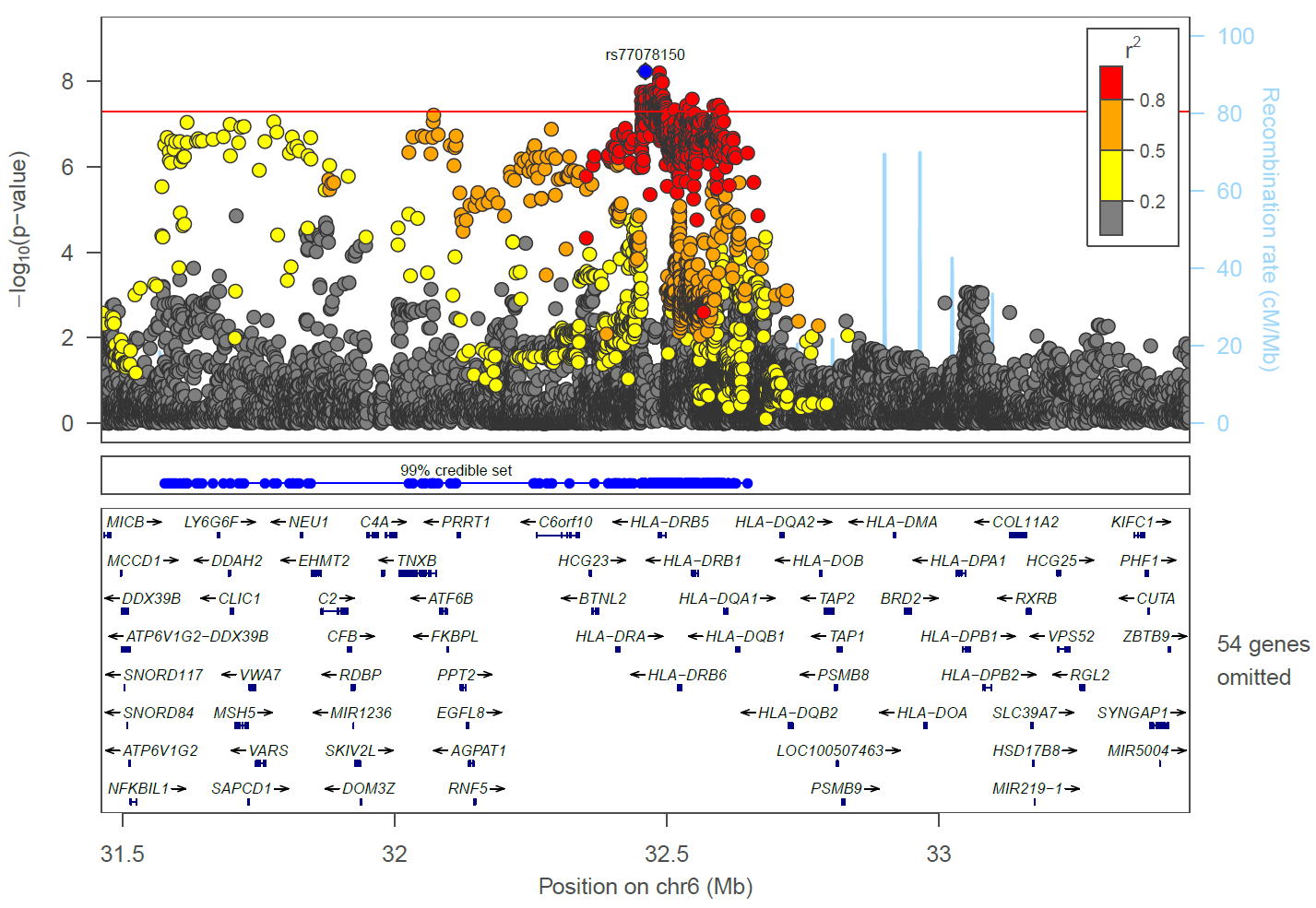

Supplementary Figure 9**:** Region plot for P2200.1_EUR central neuropathic pain *HLA-DRB5* locus, sentinel SNP labelled with a blue diamond, credible set position shown by blue dots below the region plot.

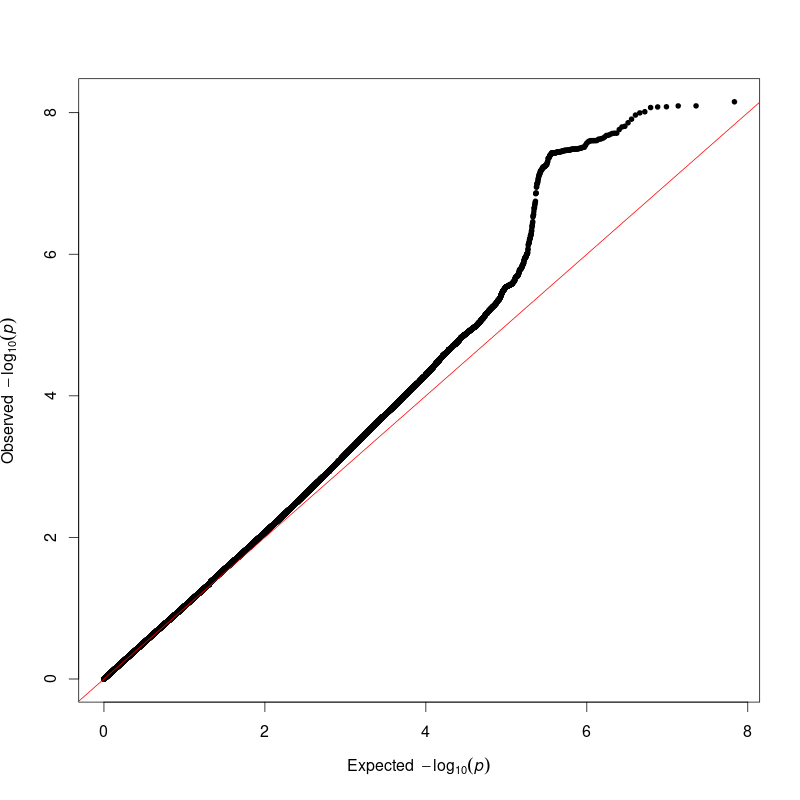

Supplementary Figure 10: QQ plot of GWAS results for P2200.3_EUR (Peripheral neuropathic pain), LD intercept 1.01

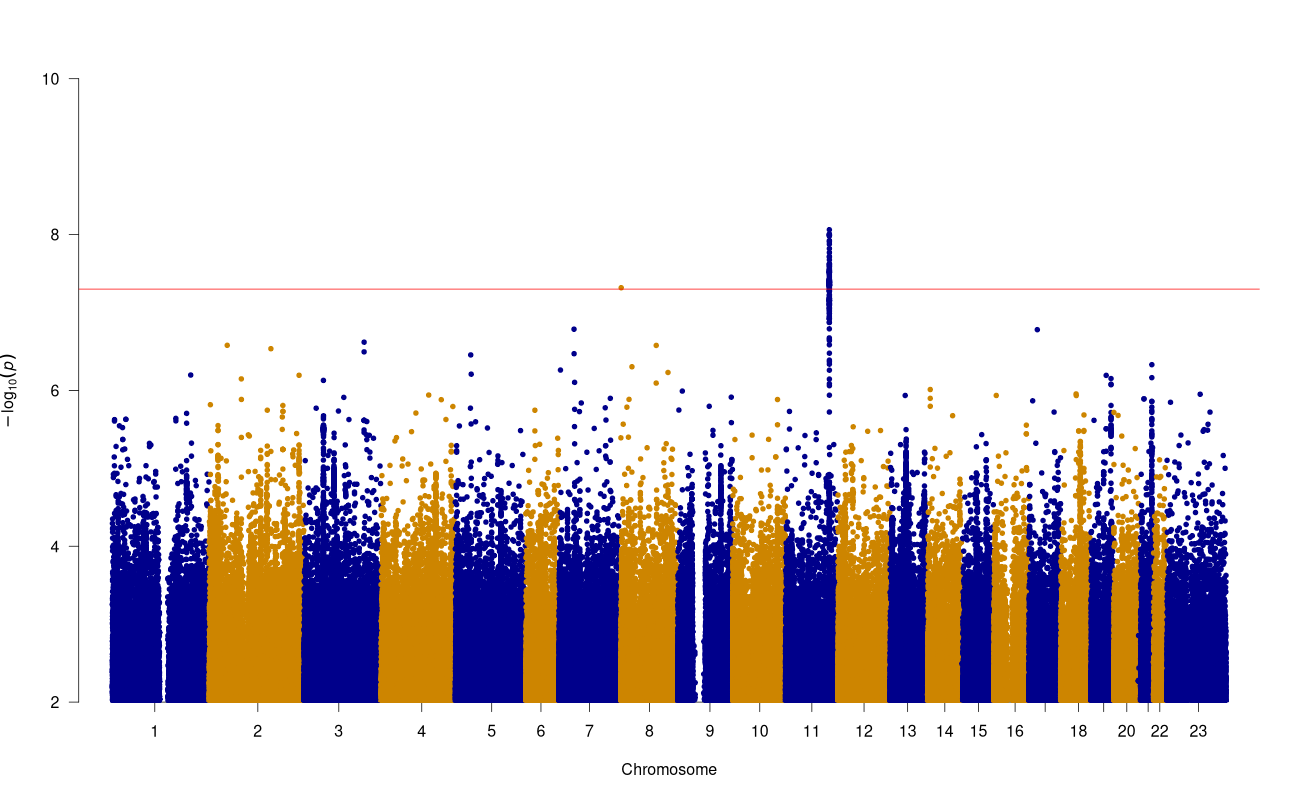

Supplementary Figure 11: Manhattan plot of GWAS results for P2200.3_EUR (Peripheral neuropathic pain) (red line=5x10^-8^, blue line=5x10^-7^)

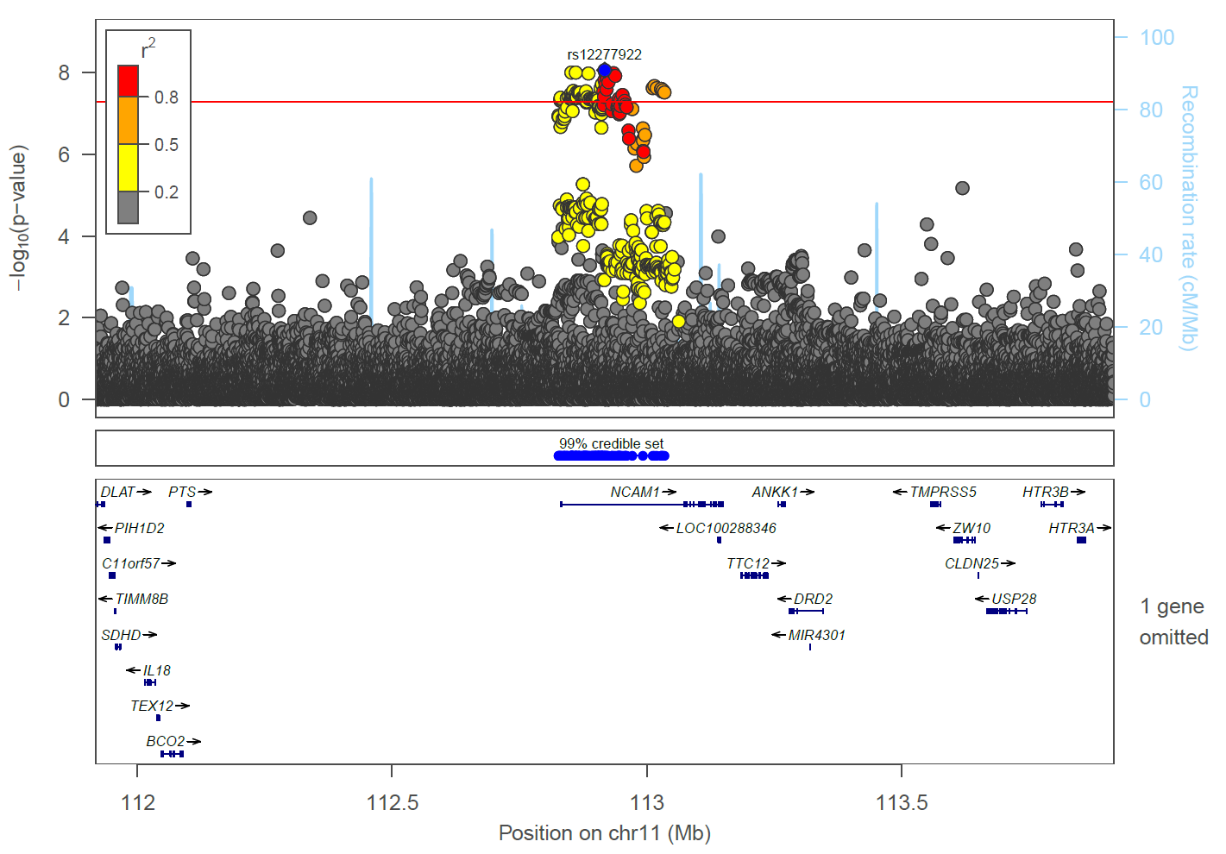

Supplementary Figure 12: Region plot for P2200.3_EUR peripheral neuropathic pain *NCAM1* locus, sentinel SNP labelled with a blue diamond, credible set position shown by blue dots below the region plot.

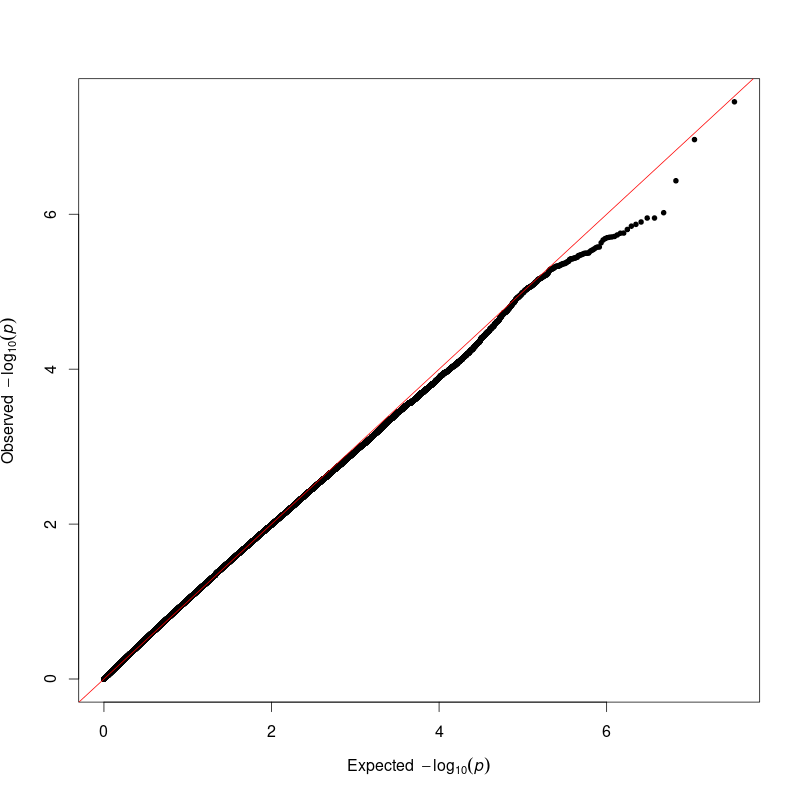

Supplementary Figure 13: QQ plot of GWAS results for P2200.3_AFR (Peripheral neuropathic pain), LD intercept 1.02

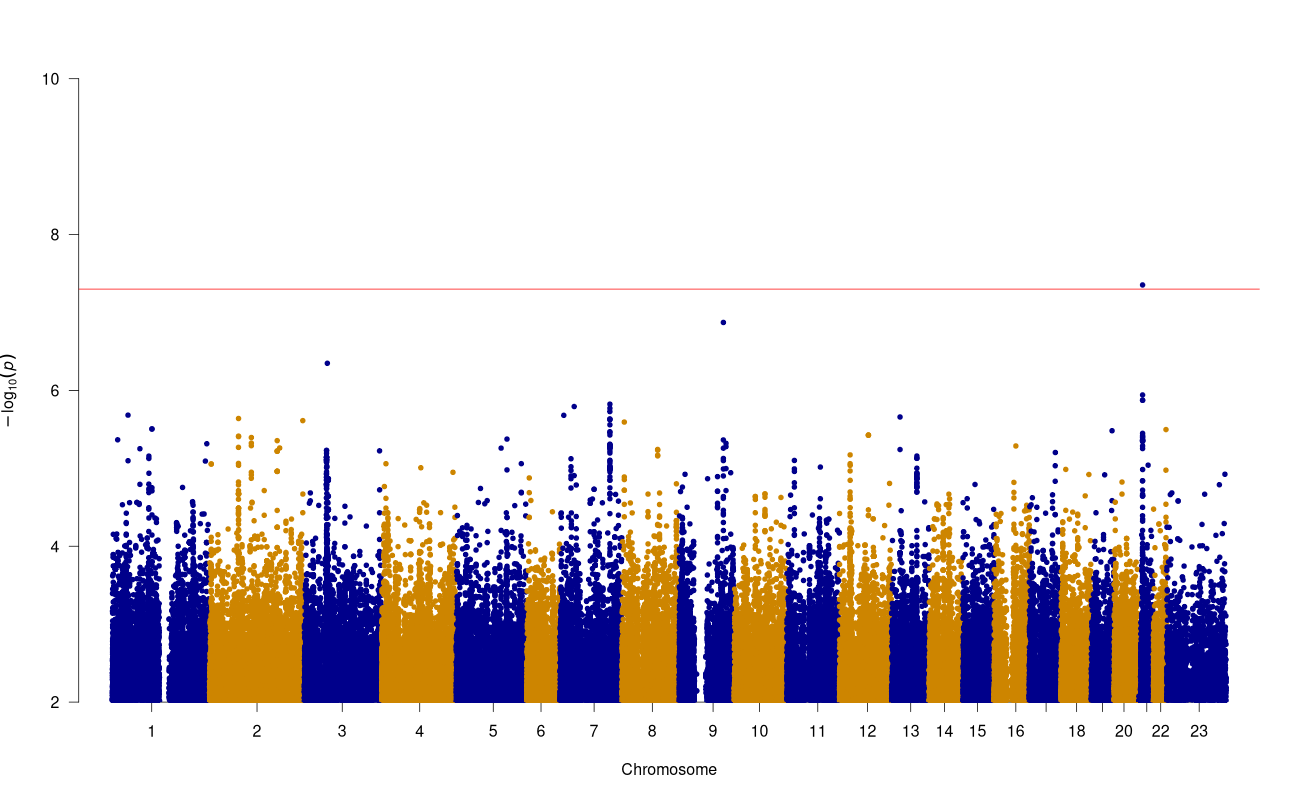

Supplementary Figure 14: Manhattan plot of GWAS results for P2200.3_AFR (Peripheral neuropathic pain) (red line=5x10^-8^, blue line=5x10^-7^)

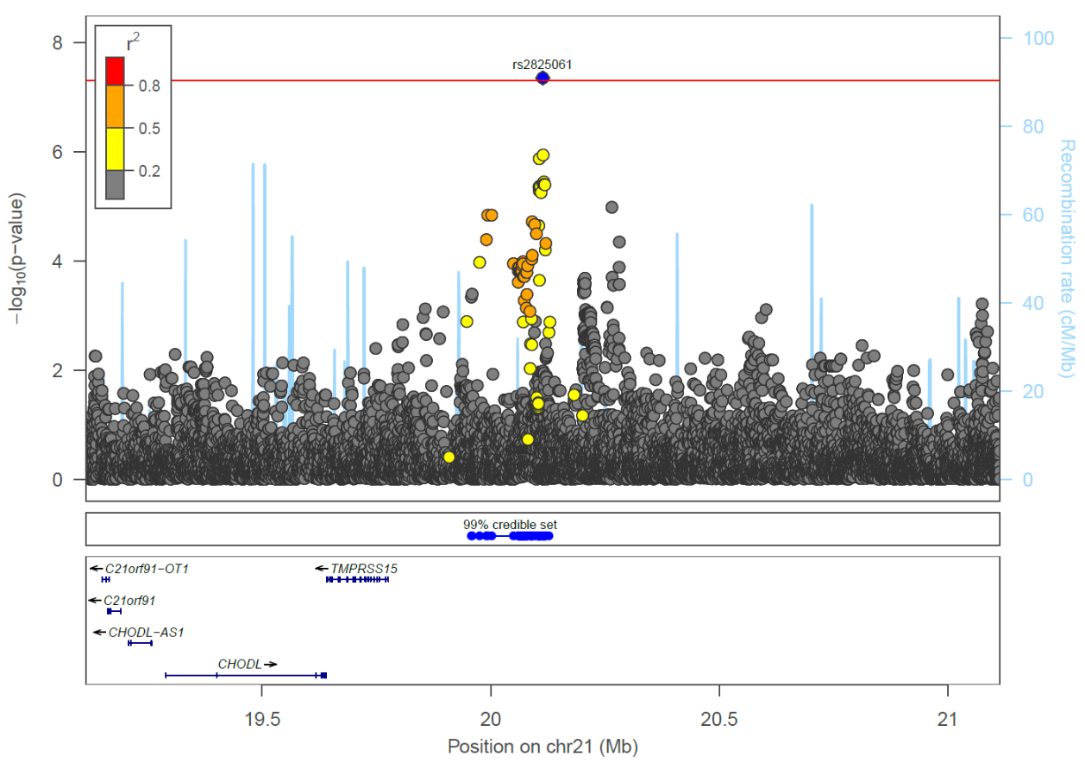

Supplementary Figure 15: Region plot for P2200.3_AFR peripheral neuropathic pain *MIR548XHG* locus, sentinel SNP labelled with a blue diamond, credible set position shown by blue dots below the region plot.

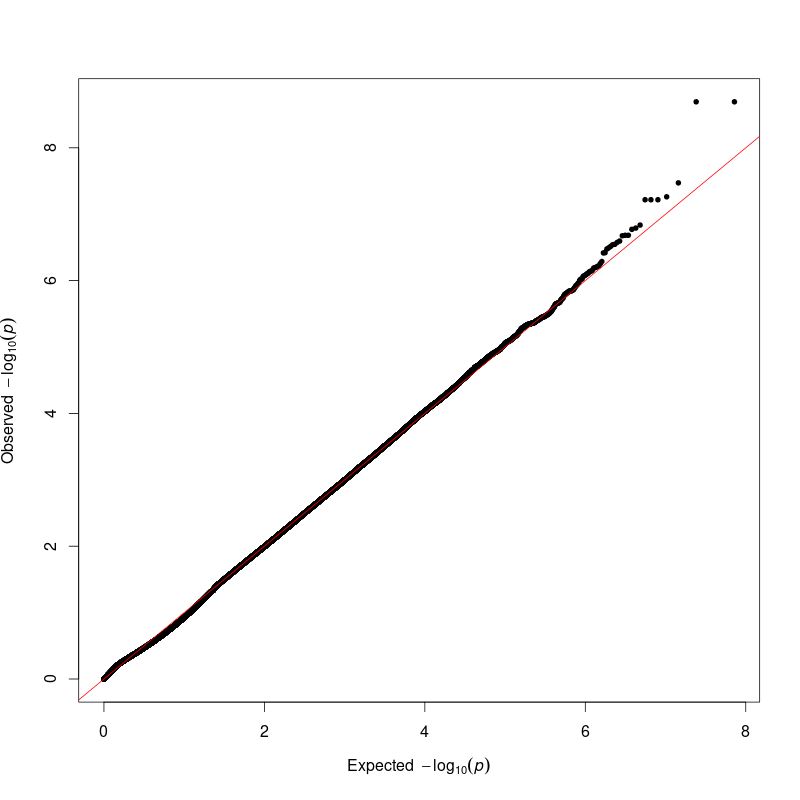

Supplementary Figure 16: QQ plot of GWAS results for P2200.39_EUR (Fibromyalgia pain clinical), LD intercept 0.99

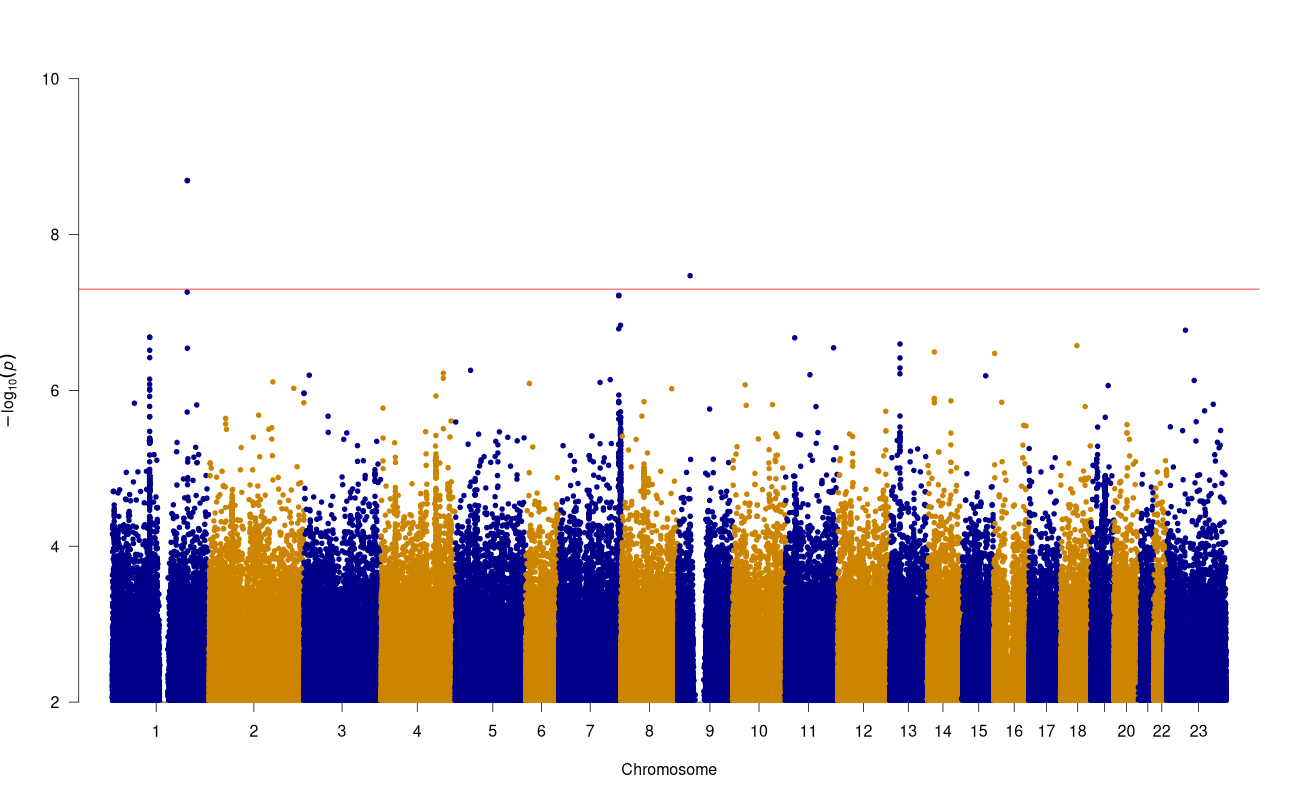

Supplementary Figure 17: Manhattan plot of GWAS results for P2200.39_EUR (Fibromyalgia pain clinical) (red line=5x10^-8^, blue line=5x10^-7^)

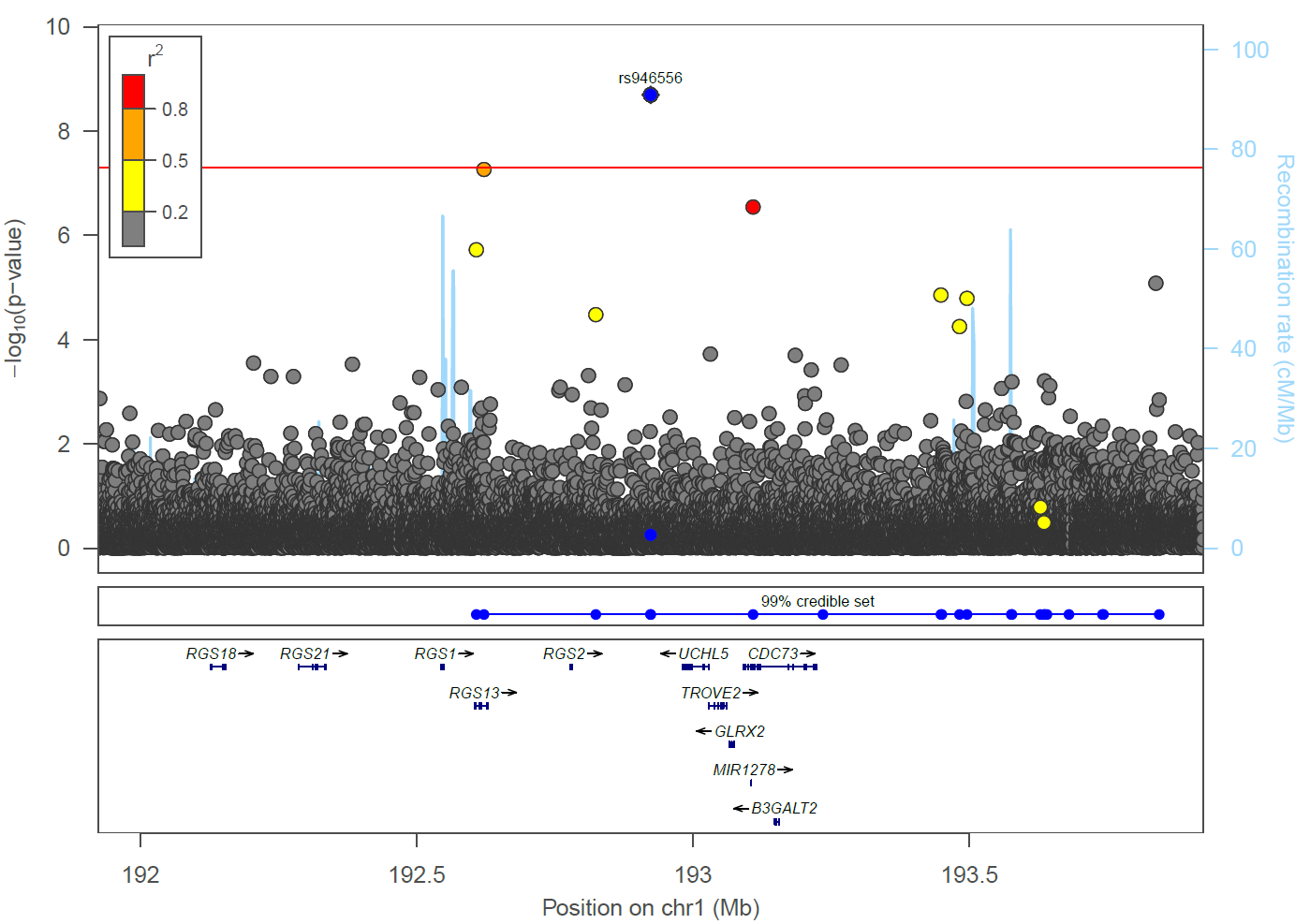

Supplementary Figure 18: Region plot for P2200.39_EUR fibromyalgia *RGS2-AS1* locus, sentinel SNP labelled with a blue diamond, credible set position shown by blue dots below the region plot.

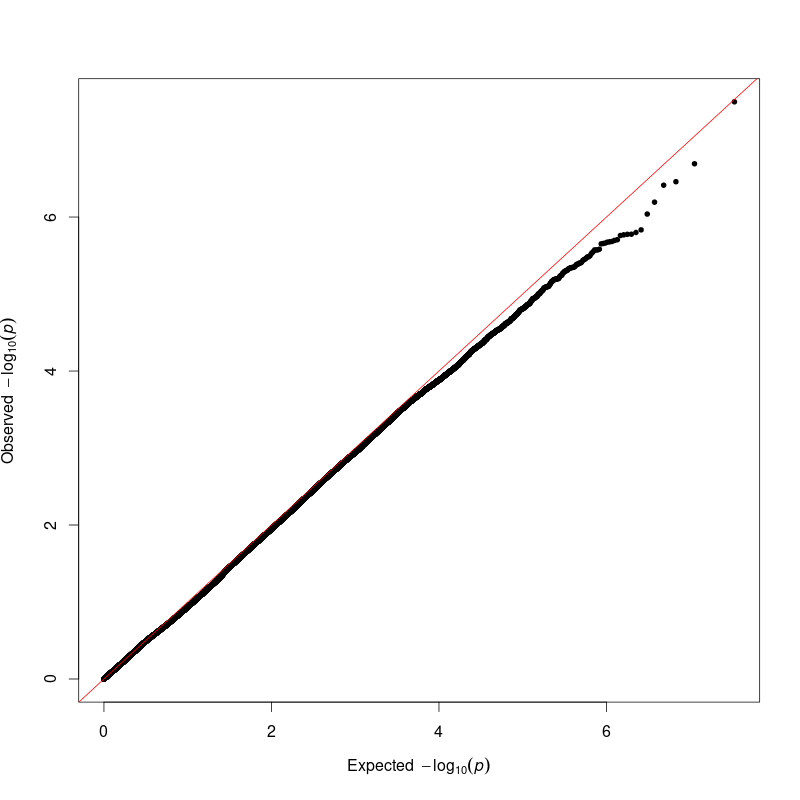

Supplementary Figure 19: QQ plot of GWAS results for P2200.40_EUR (Post-surgical neuropathic pain), LD intercept 0.98

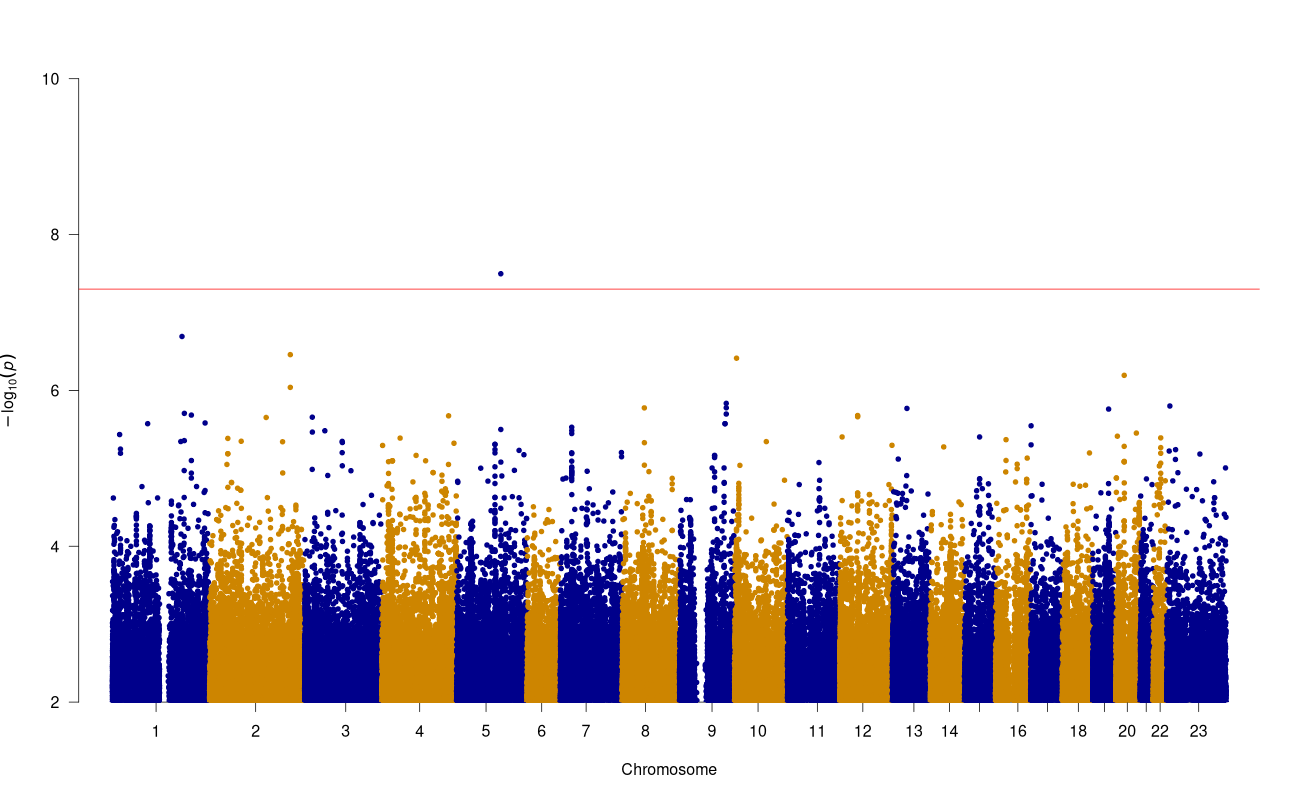

Supplementary Figure 20: Manhattan plot of GWAS results for P2200.40_EUR (Post-surgical neuropathic pain) (red line=5x10^-8^, blue line=5x10^-7^)

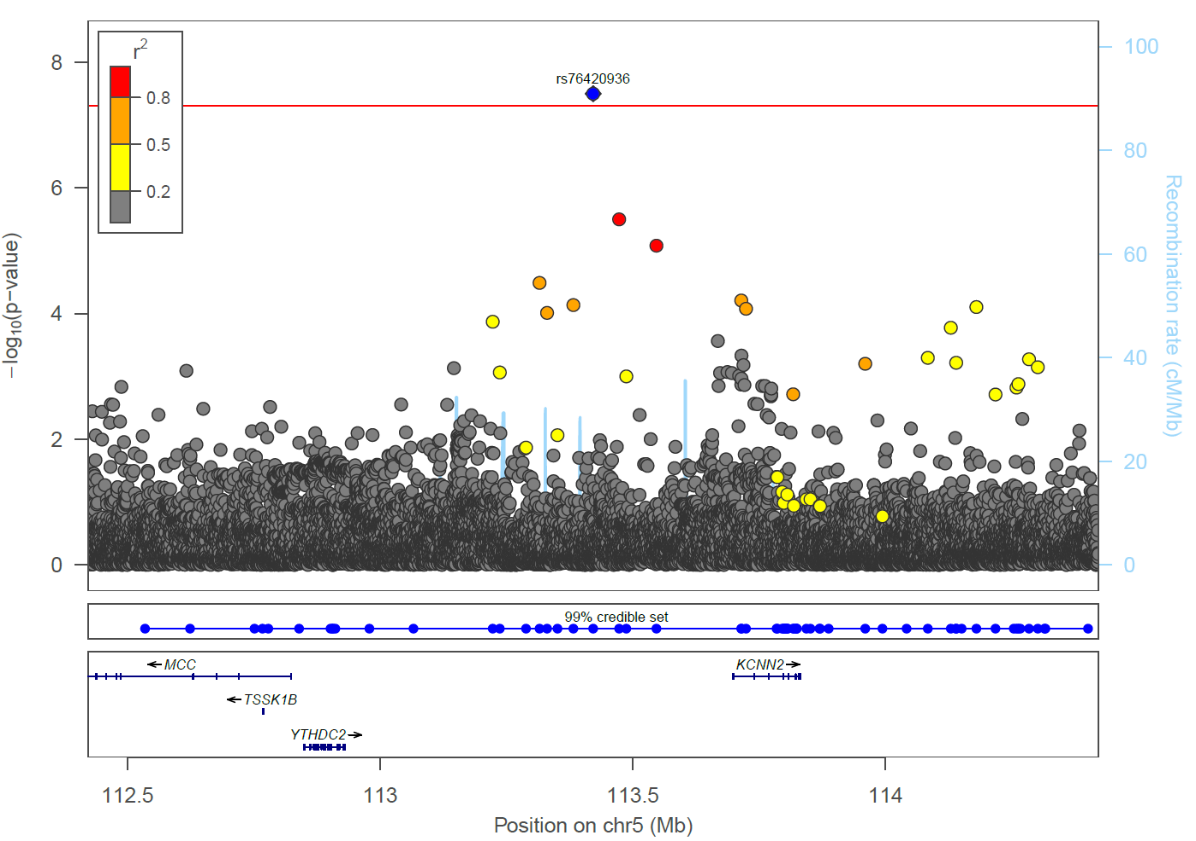

Supplementary Figure 21: Region plot for P2200.40_EUR post-surgical neuropathic pain *KCNN2* locus, sentinel SNP labelled with a blue diamond, credible set position shown by blue dots below the region plot.

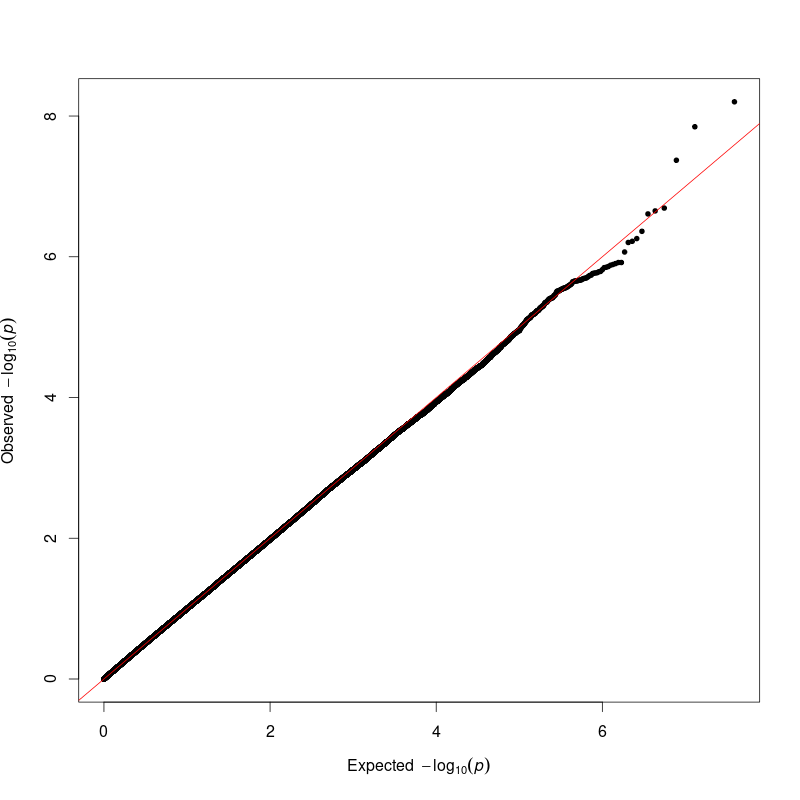

Supplementary Figure 22: QQ plot of GWAS results for P2200.511_EUR (Sciatica clinical + drugs), LD intercept 1.01

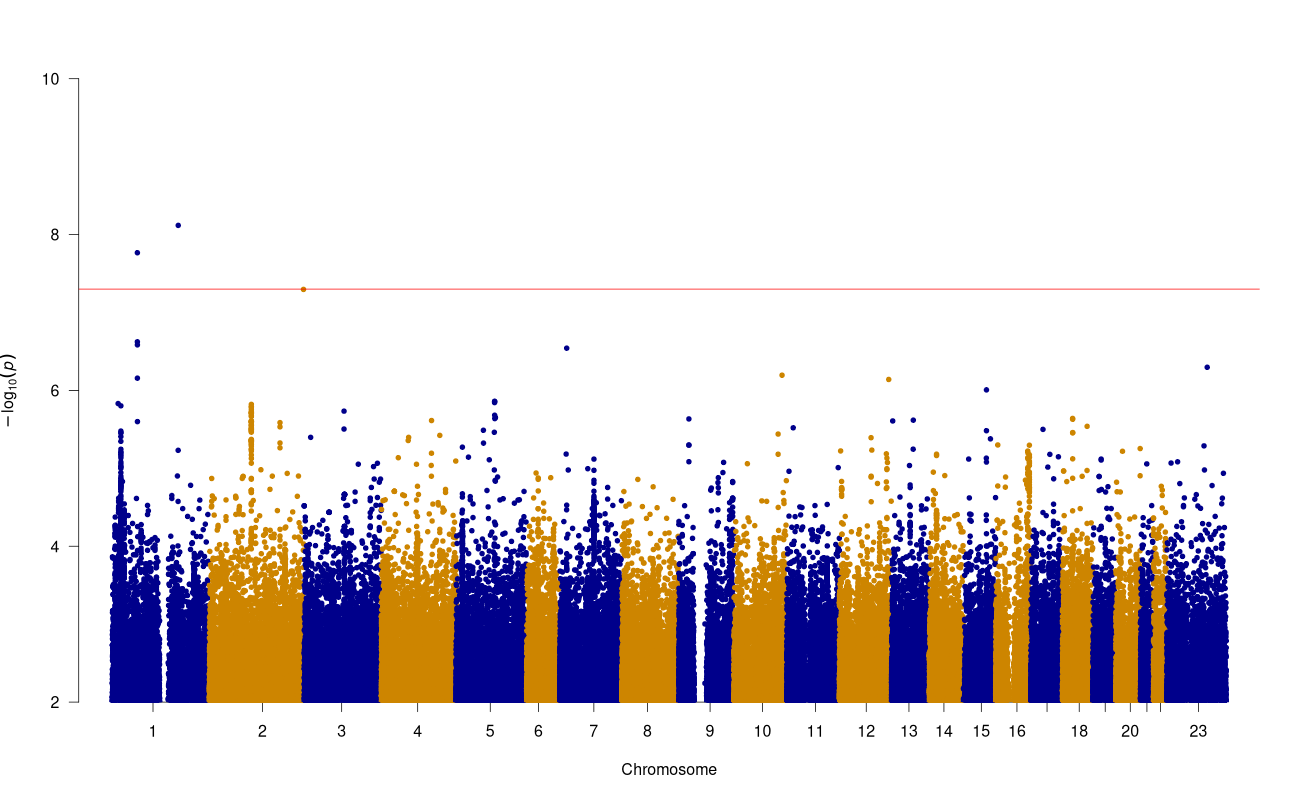

Supplementary Figure 23: Manhattan plot of GWAS results for P2200.511_EUR (Sciatica clinical + drugs) (red line=5x10^-8^, blue line=5x10^-7^)

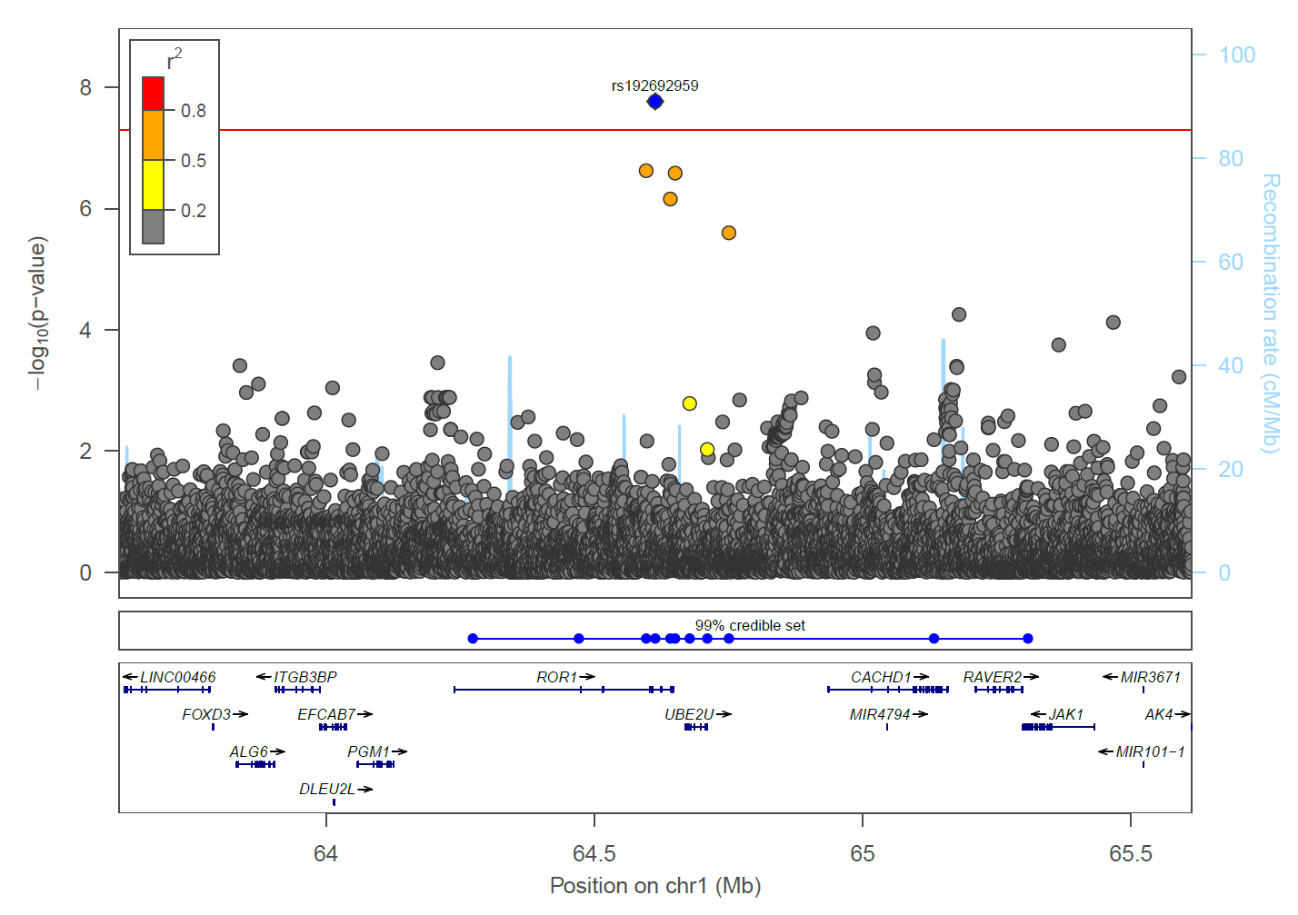

Supplementary Figure 24: Region plot for P2200.511_EUR sciatica + drugs *ROR1* locus, sentinel SNP labelled with a blue diamond, credible set position shown by blue dots below the region plot.

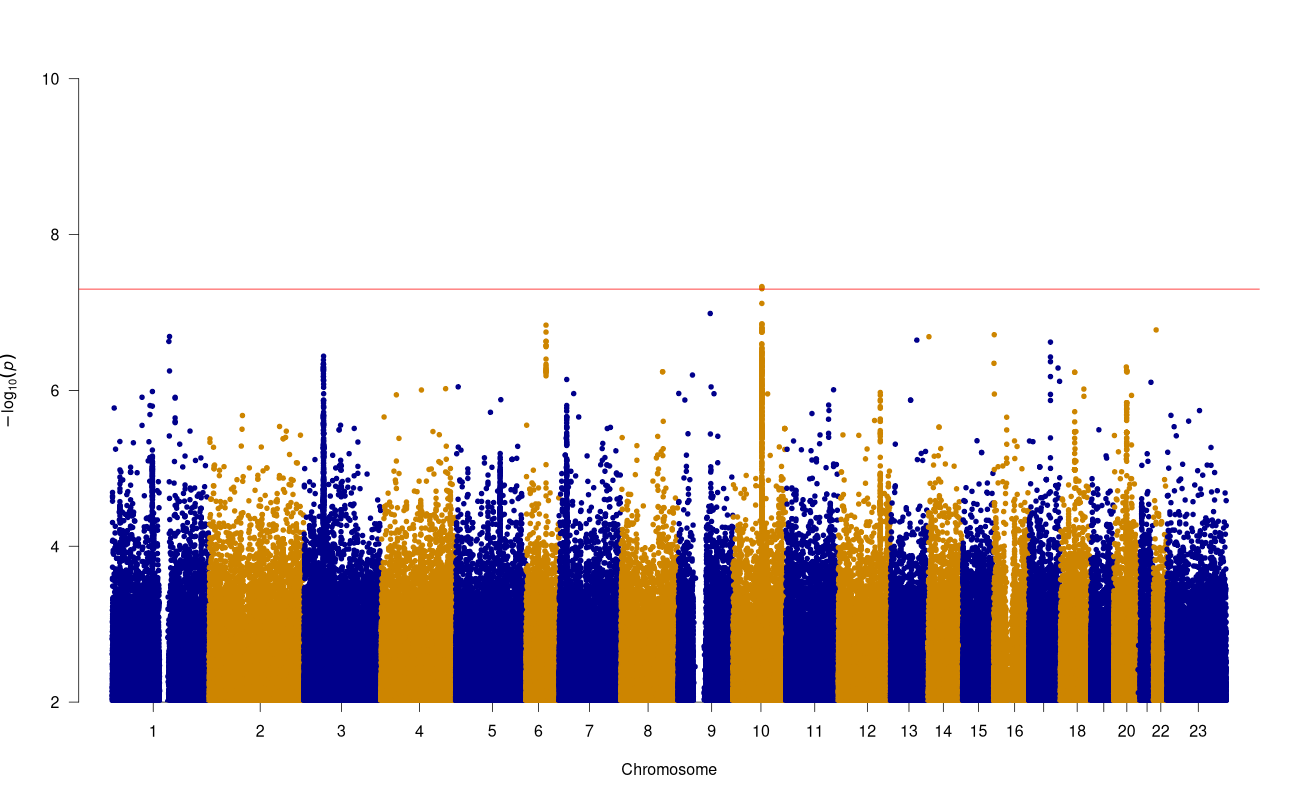

Supplementary Figure 25: Manhattan plot of GWAS results for P2200.51_EUR (Sciatica clinical) (red line=5x10^-8^, blue line=5x10^-7^)

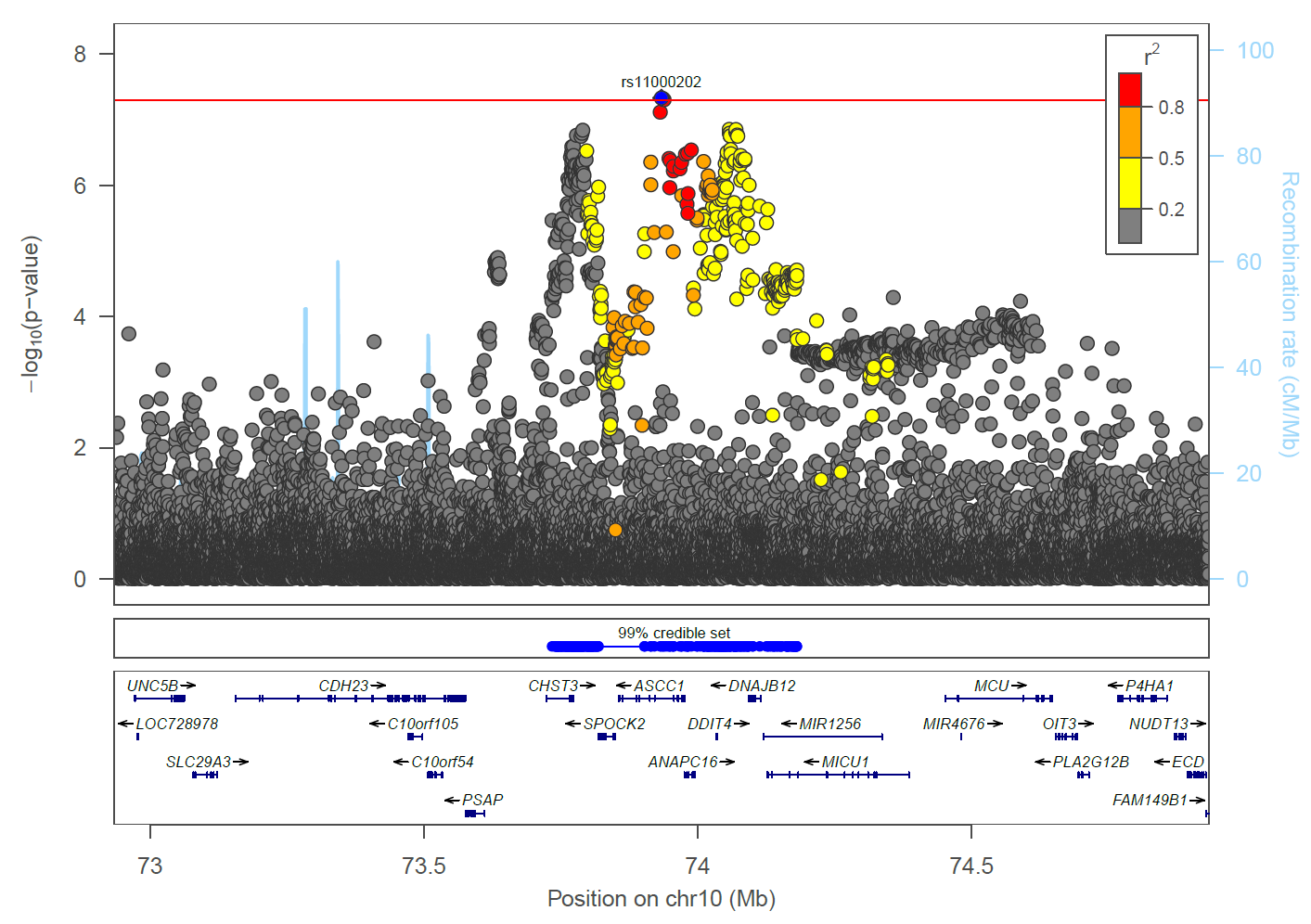

Supplementary Figure 26: Region plot for P2200.51_EUR clinical sciatica *ASCC1* locus, sentinel SNP labelled with a blue diamond, credible set position shown by blue dots below the region plot.
